# Genetics of Myocardial Interstitial Fibrosis in the Human Heart and Association with Disease

**DOI:** 10.1101/2021.11.05.21265953

**Authors:** Victor Nauffal, Paolo Di Achille, Marcus. D. R. Klarqvist, Jonathan W. Cunningham, James P. Pirruccello, Lu-Chen Weng, Valerie N. Morrill, Seung Hoan Choi, Shaan Khurshid, Samuel F. Friedman, Mahan Nekoui, Carolina Roselli, Kenney Ng, Anthony A. Philippakis, Puneet Batra, Patrick T. Ellinor, Steven A. Lubitz

**Author notes:** These authors contributed equally to this work (Victor Nauffal, Paolo Di Achille, Marcus. D. R. Klarqvist, Jonathan W. Cunningham). Jointly supervised this work (Patrick T. Ellinor, Steven A. Lubitz). **Please address correspondence to: Patrick T. Ellinor, MD, PhD, Cardiovascular Disease Initiative**, The Broad Institute of MIT and Harvard, Cambridge, MA 02124, **Steven A. Lubitz, MD, MPH, Cardiovascular Disease Initiative**, The Broad Institute of MIT and Harvard, Cambridge, MA 02124.

## Abstract

Myocardial interstitial fibrosis is a common thread in multiple cardiovascular diseases including heart failure, atrial fibrillation, conduction disease and sudden cardiac death. To investigate the biologic pathways that underlie interstitial fibrosis in the human heart, we developed a machine learning model to measure myocardial T1 time, a marker of myocardial interstitial fibrosis, in 42,654 UK Biobank participants. Greater T1 time was associated with impaired glucose metabolism, systemic inflammation, renal disease, aortic stenosis, cardiomyopathy, heart failure, atrial fibrillation and conduction disease. In genome-wide association analysis, we identified 12 independent loci associated with native myocardial T1 time with evidence of high genetic correlation between the interventricular septum and left ventricle free wall (r2g = 0.82). The identified loci implicated genes involved in glucose homeostasis (*SLC2A12*), iron homeostasis (*HFE, TMPRSS6*), tissue repair (*ADAMTSL1, VEGFC*), oxidative stress (*SOD2*), cardiac hypertrophy (*MYH7B*) and calcium signaling (*CAMK2D*). Transcriptome-wide association studies highlighted the role of expression of *ADAMTSL1* and *SLC2A12* in human cardiac tissue in modulating myocardial tissue characteristics and interstitial fibrosis. Harnessing machine learning to perform large-scale phenotyping of interstitial fibrosis in the human heart, our results yield novel insights into biologically relevant pathways for myocardial fibrosis and prioritize investigation of pathways for the development of anti-fibrotic therapies.

## INTRODUCTION

The cardiac extracellular matrix (ECM) is a dynamic compartment that plays key structural and regulatory roles in establishing myocardial tissue architecture and function. Pathologic perturbations to homeostatic turnover of ECM components leads to the progressive development of interstitial fibrosis,^1^ which is the histological hallmark of several cardiac diseases including cardiomyopathy,^2^ heart failure,^3,4^ valvular heart disease,^5^ conduction disease,^6^ atrial fibrillation,^7^ and sudden cardiac death.^8^ Interstitial fibrosis is commonplace in the aging heart,^9,10^ yet a myriad of hemodynamic, metabolic and inflammatory stressors contribute to the accelerated development of interstitial fibrosis and associated cardiovascular diseases.^11^ There is a critical need to understand the biological basis and mechanisms of cardiac fibrosis in humans, since identification of the mechanisms of fibrosis may enable opportunities to prevent the process and could have a wide-ranging impact on multiple cardiovascular diseases. However, progress has been hindered by challenges in reliable non-invasive measurement of interstitial fibrosis at scale in the human heart, and by lack of adequately powered validation studies of findings from animal or tissue/cell models in humans. The advent of machine learning tools capable of generation of imaging-based phenotypes at scale and large biorepositories with deep phenotyping and genomic data, offers a unique opportunity to overcome these challenges.

Native myocardial T1 time measured using cardiac magnetic resonance imaging (cMRI) is a histopathologically validated metric for quantifying interstitial fibrosis in the human heart.^12,13^ The UK Biobank is a large-scale prospective cohort with rich cardiac magnetic resonance imaging^14^, genomic, and clinical outcomes data.^15^ We sought to use machine learning to quantify fibrosis in over 40,000 study participants who underwent cMRI T1 mapping, assess associations between fibrosis and clinical outcomes, and identify pathways responsible for cardiac fibrosis in humans using genetic association analyses.

## RESULTS

### Machine Learning-Derived T1 Time is Highly Correlated with Manually-Derived T1 Time and Enables T1 Time Measurement at Scale

We acquired mid-ventricular, short-axis cMRI T1 maps for 42,654 participants in the UK Biobank (**Figure 1**). The mean age of the participants was 64.1 ± 7.7 years and 48% were men (**Table 1**). We first selected 600 random T1 maps (500 for training and 100 for validation) to develop our machine learning model (**Supplemental Figure 1**). To train a machine learning model to segment the target cardiac structures of interest (interventricular septum (IVS) and left ventricle free wall (LV FW)), two cardiologists manually traced and labelled the selected structures of interest in the 500 training images (**Supplemental Figure 2** and see **Online Methods**). The manual tracing procedure, called semantic segmentation, displayed high inter-reader concordance between the cardiologist-labelled segmentations, as measured in 50 overlapping cMRI acquisitions (Sørensen-Dice coefficients: IVS 0.84, 95% CI 0.76 - 0.92; LV FW 0.79, 95% CI 0.63 - 0.95). Additionally, T1 derived times were highly correlated between the two readers (Pearson correlation coefficient r: IVS 0.95, 95% CI 0.92 - 0.97; LV FW 0.84, 95% CI 0.73 - 0.91) (**Supplemental Figure 3**).

**Table 1.** Study sample characteristics at time of first visit for cardiac magnetic resonance imaging.

| Baseline characteristic | N (%) |
| --- | --- |
| Participants | 42,654 |
| Age at MRI (mean (SD)) | 64.1 (7.7) |
| Male | 20,493 (48.0) |
| Body mass index, kg/m<sup>2</sup> (mean (SD)) | 26.47 (4.35) |
| Current cigarette smoker | 1,459 (3.4) |
| Hypertension | 12,983 (30.4) |
| Diabetes mellitus type 2 | 1,579 (3.7) |
| Diabetes mellitus type 1 | 167 (0.4) |
| Hyperlipidemia | 445 (1.0) |
| Chronic kidney disease | 360 (0.8) |
| Coronary artery disease | 2,546 (6.0) |
| Myocardial infarction | 875 (2.1) |
| Dilated cardiomyopathy | 90 (0.2) |
| Hypertrophic cardiomyopathy | 31 (0.1) |
| Heart failure | 287 (0.7) |
| Aortic stenosis | 80 (0.2) |
| Pulmonary hypertension | 20 (0.0) |
| Atrial fibrillation | 1,241 (2.9) |
| Ventricular arrhythmia/Cardiac arrest | 156 (0.4) |
| Atrioventricular/Distal conduction disease | 294 (0.7) |
| Rheumatoid Arthritis | 603 (1.4) |
| Beta blocker | 2,305 (5.4) |
| ACE-inhibitor/ARB | 5,912 (13.9) |
| Mineralocorticoid receptor antagonist | 81 (0.2) |
| Statins | 8,278 (19.4) |
| Interventricular septum T1 time, ms (mean (SD)) | 918.1 (41.5) |
| LV free wall T1 time, ms (mean (SD)) | 902.1 (45.0) |
| LV mass, g (mean (SD)) | 90.1 (29.7) |
| LV ejection fraction, % (mean (SD)) | 60 (10) |
| LV end systolic volume, ml (mean (SD)) | 58.1 (19.0) |
| LV end diastolic volume, ml (mean (SD)) | 143.2 (32.8) |
| LV stroke volume, ml (mean (SD)) | 85.2 (18.5) |
| LA end systolic volume, ml (mean (SD)) | 33.6 (14.2) |
| LA end diastolic volume, ml (mean (SD)) | 72.8 (20.3) |
| LA ejection fraction, % (mean (SD)) | 54.7 (8.6) |

Values are presented as the number (percentage) unless otherwise specified. A subset of study participants had data available for each cMRI parameter: Interventricular septum T1 time (n=41,505); left ventricle free wall T1 time (n=39,311); LA end systolic volume/LA end diastolic volume/LA ejection fraction (n=38,261); LV end systolic volume/LV end diastolic volume/LV ejection fraction (n=42,021); LV mass (n=41,900). ACE: angiotensin converting enzyme, ARB: angiotensin receptor blocker, SD: standard deviation, LA: left atrium, LV: left ventricle.

**Figure 1.**
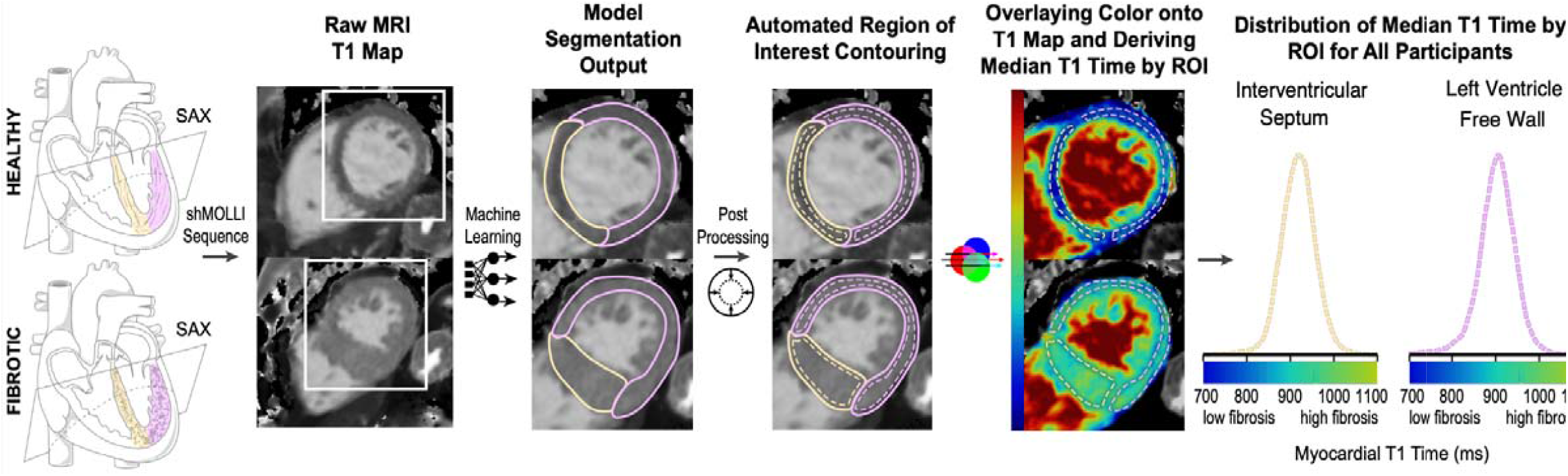
Overview of the automated pipeline for native myocardial T1 time measurement at the interventricular septum and left ventricle free wall using machine learning. A representative healthy heart and one with increased interstitial fibrosis are shown for illustration. Cardiac T1 mapping using the Shortened Modified Look-Locker Inversion (shMOLLI) recovery sequence was performed at the mid-ventricular short-axis. A machine-learning model trained on the raw MRI T1 maps generated automated segmentation of the interventricular segment and left ventricle free wall followed by selection of representative myocardial regions of interest using morphological erosion. T1 map color legends were then used to transform pixel intensities within the regions of interest into T1 times. For each participant the median T1 time by ROI was calculated and used as the representative T1 time for that segment. ROI: regions of interest; shMOLLI: Shortened Modified Look-Locker Inversion.

We then trained a machine learning model to identify the IVS and the LV FW using the cardiologist-segmented data as truth labels. The machine learning model had high accuracy when tested in the validation set (Sørensen-Dice coefficients: IVS 0.82, 95% CI 0.70 - 0.94; LV FW 0.81, 95% CI 0.67 - 0.95). Model predictions were then post-processed using morphological erosion of the segments of interest to automatically generate representative mid-myocardial regions of interest within the IVS and LV FW. Native T1 time for each segment was computed using the median pixel intensity, converted to T1 time, in the respective region of interest (**Figure 1**). In the validation set, automatically generated T1 times were highly correlated with T1 times derived from manually-traced regions of interest (Pearson correlation coefficient r: IVS 0.97, 95% CI 0.95 - 0.98; LV FW 0.92, 95% CI 0.89 - 0.95) (**Supplemental Figure 4**).

Next, the machine learning model was used to segment the IVS and LV FW in the remaining 42,054 cMRIs not used for model training or validation followed by automated selection of regions of interest and measurement of native T1 time. To maximize the quality of the generated native myocardial T1 times, we manually reviewed all 42,654 T1 maps to exclude low-quality acquisitions and major artifacts within our segments of interest (see **Online Methods**). Following quality control, we were able to measure native myocardial T1 time for the IVS and LV FW in 41,505 and 39,311 individuals, respectively (**Supplemental Figure 1**). We found that the LV FW segment had a 3-fold higher incidence of artifact as compared to the interventricular septum (N _LV FW Major Artifact_ = 3,343 vs. N _IVS Major Artifact_ = 1,149). The artifact-prone nature of the LV FW segment is well recognized in the field of cardiac T1 mapping and has led some experts to advocate for preferential use of interventricular septum T1 time to differentiate health and disease states of the myocardium.^16,17^ As such, while we present results for both the IVS and LV FW, the IVS native myocardial T1 time constituted our primary analysis.

Mean IVS and LV FW T1 time of the study sample was 918.1 ± 41.5 ms and 902.1 ± 45.0 ms, respectively. These values are consistent with previously reported T1 times in a smaller study from the UK Biobank^18^ including 11,882 cMRIs and other population-based studies with cardiac T1 mapping using 1.5 T MRI scanners.^10^ Furthermore, our results are consistent with known gender-specific patterns of higher native myocardial T1 time in women compared to men (**Supplemental Figure 5**).^10,19^ There was moderate correlation between IVS and LV FW T1 time (Pearson Correlation Coefficient r: IVS 0.64, 95% CI 0.63 - 0.65; **Supplemental Figure 6**) in the study sample.

### Native Myocardial T1 Time is Associated with Prevalent Cardiovascular, Metabolic and Systemic Inflammatory Diseases

We then investigated if native myocardial T1 times are associated with cardiovascular, metabolic and systemic inflammatory diseases by comparing the native T1 times of the IVS and LV FW from participants with prevalent disease at the time of cardiac MRI to healthy participants. Healthy participants were selected to be free of prevalent dilated cardiomyopathy, hypertrophic cardiomyopathy, heart failure, atrial fibrillation, atrioventricular node/distal conduction disease, hypertension, diabetes mellitus, aortic stenosis, chronic kidney disease, hemochromatosis and rheumatoid arthritis. In multivariable analyses, we found that multiple cardiovascular, metabolic and systemic inflammatory diseases were associated with increased native myocardial T1 time of the IVS and LV FW segments (**Figure 2** and **Supplemental Figure 7**). Directionality of the associations for both segments was generally consistent, albeit with varying effect sizes and significance levels for each segment.

**Figure 2.**
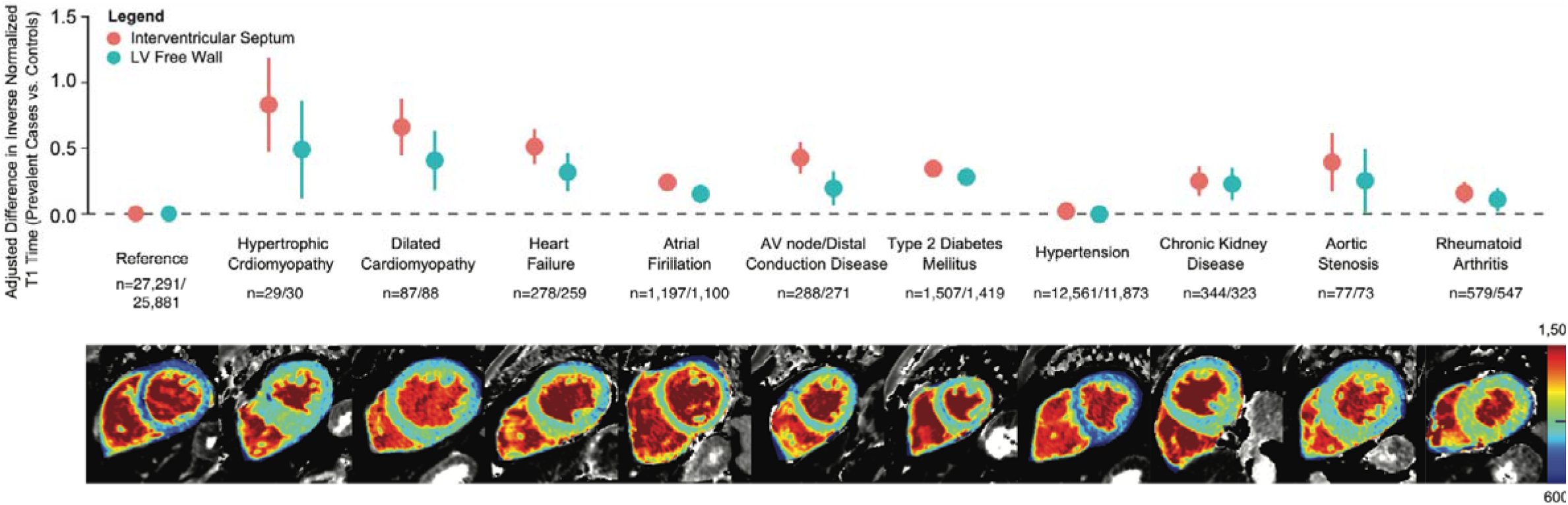
Change in native myocardial T1 time associated with prevalent cardiovascular, metabolic and systemic inflammatory diseases as compared to healthy controls. Healthy controls free of prevalent dilated cardiomyopathy, hypertrophic cardiomyopathy, heart failure, atrial fibrillation, atrioventricular node/distal conduction disease, hypertension, diabetes mellitus, aortic stenosis, chronic kidney disease, hemochromatosis and rheumatoid arthritis constituted the reference group. Numbers of controls or cases with available IVS/LV FW T1 time are shown below each category. For each disease a representative T1 map of a case is provided from the study sample. AV: atrioventricular.

Among cardiovascular diseases, higher native myocardial T1 time in both segments of interest was associated with hypertrophic cardiomyopathy (P_IVS_ = 5.8 × 10^−6^ and P_LV FW_ = 1.0 × 10^−2^), dilated cardiomyopathy (P_IVS_ = 1.6 × 10^−9^ and P_LV FW_ = 4.1 × 10^−4^), heart failure (P_IVS_ = 9.8 × 10^−14^ and P_LV FW_ = 2.1 × 10^−5^), atrial fibrillation (P_IVS_ = 1.6 × 10^−12^ and P_LV FW_ = 3.9 ×10^−5^), atrioventricular node/distal conduction disease (P_IVS_ = 2.9 × 10^−12^ and P_LV FW_ = 3.1 × 10^−3^) and, aortic stenosis (P_IVS_ = 5.5 × 10^−4^ and P_LV FW_ = 4.0 × 10^−2^). For coronary artery disease (P_IVS_ = 8.0 × 10^−3^ and P_LV FW_ = 0.25), myocardial infarction (P_IVS_ = 6.8 × 10^−3^ and P_LV FW_ = 0.26), and history of cardiac arrest (P_IVS_ = 3.4 × 10^−2^ and P_LV FW_ = 0.05), only higher native T1 times of the IVS was associated with disease. Native T1 time was not associated with hypertension (P_IVS_ = 0.16 and P_LV FW_ = 0.79) which is consistent with prior studies that showed limited ability of T1 mapping to differentiate between individuals with hypertension and controls except among individuals with inadequately controlled hypertension and concomitant left ventricular hypertrophy (**Figure 2** and **Supplemental Figure 7)**.^20^

Metabolic disorders, including type-2 diabetes mellitus (P_IVS_ = 6.3 × 10^−27^ and P_LV FW_ = 5.3 × 10^− 16^), type-1 diabetes mellitus (P_IVS_ = 1.5 × 10^−8^ and P_LV FW_ = 3.5 × 10^−3^), hyperlipidemia (P_IVS_ = 5.5 × 10^−5^ and P_LV FW_ = 4.8 × 10^−2^) and chronic kidney disease (P_IVS_ = 1.7 × 10^−5^ and P_LV FW_ = 4.0 × 10^−2^) were associated with significantly higher native T1 times in both segments (**Figure 2** and **Supplemental Figure 7**).

Of the systemic inflammatory diseases examined, rheumatoid arthritis was associated with significantly increased native T1 time of both segments (P_IVS_ = 1.6 × 10^−4^ and P_LV FW_ = 1.8 × 10^−2^). Notably, in the subset of participants in whom C-reactive protein was measured at enrollment (n_IVS_ = 38,731, n_LV FW_ = 36,663), we found significantly higher native T1 time in both segments among those in the top 20th percentile of the C-reactive protein distribution (P_IVS_ = 1.0 × 10^−7^ and P_LV FW_ = 3.7 ×10^−3^). Systemic lupus erythematosus was associated with increased myocardial native T1 time of the LV FW but not IVS (P_IVS_ = 0.50 and P_LV FW_ = 1.3 × 10^−2^) (**Figure 2** and **Supplemental Figure 7**).

In a sensitivity analysis, overall findings persisted with further adjustment for LV ejection fraction and LV mass, suggesting that myocardial tissue characterization with T1 mapping is a marker of myocardial changes associated with disease independent of these measures of LV function and structure (**Supplemental Figure 8**).

### Native Myocardial T1 Time is Associated with Incident Cardiovascular Disease

We then examined associations between native myocardial T1 time and incident cardiovascular diseases. Given the relatively short follow-up time following cMRI (median follow-up 2.54 years, interquartile range 1.63 - 3.88) and overall low event rate in the UK biobank, we focused on incident heart failure, atrial fibrillation and atrioventricular node/distal conduction disease given a relatively higher number of events. We compared individuals in the top 20th percentile of native T1 time distribution for the IVS and LV FW to the bottom 80th percentile. We found that participants in the top 20th percentile of IVS T1 time had a significantly higher risk of incident heart failure (HR 1.33, 95% CI 1.14 - 1.54), atrial fibrillation (HR_IVS_ 1.19, 95% CI 1.07 - 1.32) and atrioventricular node/distal conduction disease (HR 1.25, 95% CI 1.09 - 1.42) compared to those in the lower 80th percentile (**Figure 3 and Supplemental Figure 9**). Participants in the top 20th percentile of LV FW T1 time had a higher risk of incident heart failure (HR 1.33, 95% CI 1.13 - 1.57) and atrial fibrillation (HR 1.17, 95% CI 1.05 - 1.31) but not atrioventricular node/distal conduction disease (HR 1.06, 95% CI 0.92 - 1.22) compared to those in the lower 80th percentile (**Supplemental Figure 9**).

**Figure 3.**
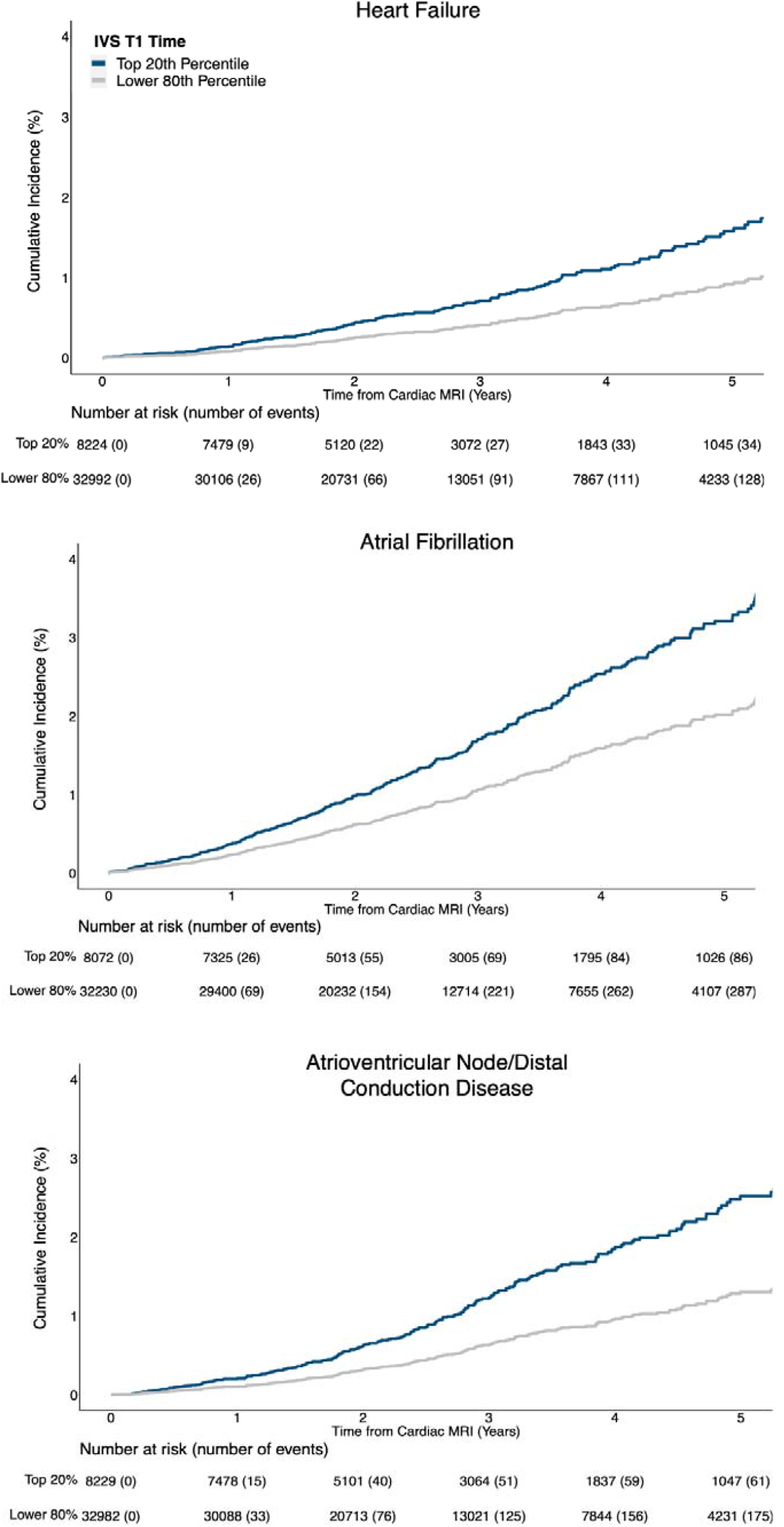
Adjusted cumulative incidence of heart failure, atrial fibrillation and atrioventricular node/distal conduction disease stratified by top 20th percentile vs. lower 80th percentile of interventricular septum T1 time. IVS: interventricular septum.

### Native Myocardial T1 Time is Heritable with Distinct Genetic Architecture Compared to Other cMRI-derived Measures of Left Ventricular and Atrial Structure and Function

Next, we sought to determine the genetic basis of interstitial fibrosis by performing genetic analyses of the IVS and LV FW T1 times. The SNP-heritability of these T1 time-based phenotypes were 0.13 (h2g_IVS_) and 0.11(h2g_LV FW_). In contrast, SNP-heritability of other cMRI phenotypes such as LV mass, LV end diastolic volume, and LV end systolic volume in the UK biobank have been reported to be 0.26, 0.40, and 0.31, respectively.^21^ The genetic correlation between IVS and LV FW was high at 0.82 implying shared genetic determinants of diffuse myocardial fibrosis throughout the left ventricle. We then examined the genetic correlation between native myocardial T1 time and cMRI measures of left ventricular and left atrial structure and function. Interestingly, we observed limited genetic correlation for both IVS and LV FW T1 time with other cMRI measures suggesting that distinct biologic pathways contribute to development of myocardial interstitial fibrosis (**Supplemental Figure 10**).

### Common Variant Association Analysis of Native Myocardial T1 Time Highlight Loci Biologically Relevant to Fibrosis

After establishing the heritability of the native T1 times, we performed genome-wide association studies (GWAS) for these traits and discovered 11 genome-wide significant loci for the IVS (**Figure 4a and Supplemental Table 1**) and 5 for the LV FW (**Figure 4b and Supplemental Table 2**). There was no evidence of inflation in our GWAS results (IVS: λGC =1.053, LD score regression intercept = 1.0002; LV FW: λGC =1.044, LD score regression intercept =1.0067) (**Supplemental Figure 11**). Regional association plots for genome-wide significant SNPs are shown in **Supplemental Figures 12 and 13**.

**Figure 4.**
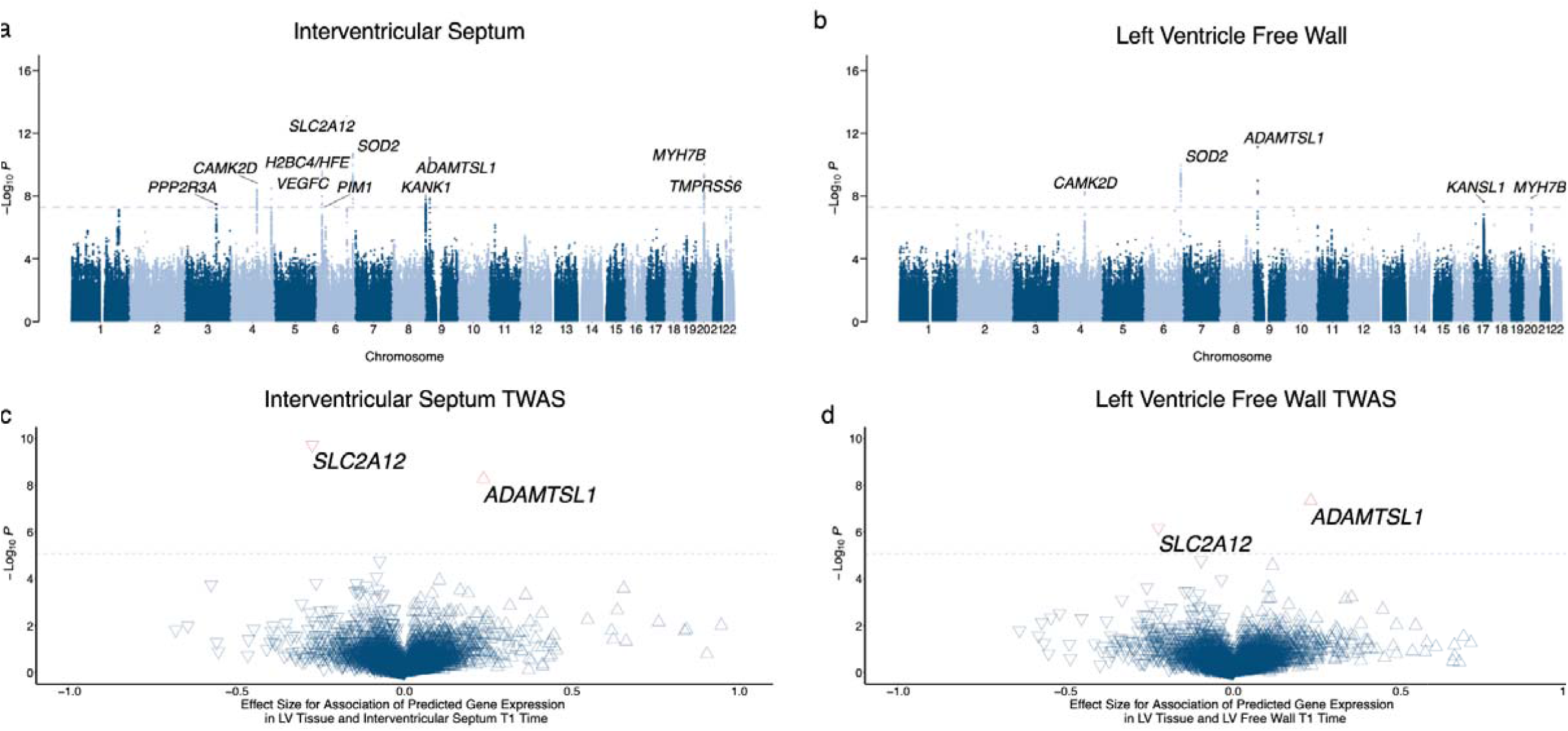
Interventricular septum (a) and left ventricle (b) native T1 time genome-wide association results across 22 autosomes. Nearest genes are used for annotation. The dashed grey line represents the threshold for genome-wide significance (P<5×10^−8^). Volcano plots depicting transcriptome-wide association results for interventricular septum (c) and LV free wall (d) native myocardial T1 time using human left ventricular tissue gene expression. Upward facing triangles reflect increased T1 time associated with increased gene expression in left ventricular tissue. Downward facing triangles reflect decreased T1 time associated with increased gene expression in left ventricular tissue.

In the IVS GWAS, a regulatory region variant in the solute carrier *SLC2A12* was the most significant lead SNP (rs2627230, P = 8.08×10^−14^) and associated with increased levels of native T1 time. *SLC2A12* encodes solute facilitated glucose transporter member 12 (GLUT12), a basal and insulin-independent glucose transporter in the heart^22^ with previously reported associations with heart failure,^23^ diabetes,^24^ and kidney disease.^25^ Lead SNPs in *SOD2* (rs6912979, P = 1.06×10^−10^) and *VEGFC* (rs365843, P = 3.19×10^−9^), two genes with established roles in cardiac hypertrophy and fibrosis in animal models,^26,27^ were associated with decreased interventricular septum T1 times. Another lead SNP associated with interventricular septum T1 time is an intronic variant in *ADAMTSL1* (rs1576900, P = 3.63×10^−11^), a gene encoding an ADAMTS-like protein which is thought to modulate the function of ADAMTS metalloproteinases with integral roles in extracellular matrix turnover.^28,29^ Additionally, *MYH7B*, a gene associated with familial hypertrophic cardiomyopathy,^30^ was among the genome-wide significant loci (rs6120777, P = 9.2×10^−11^); this association persisted even after exclusion of hypertrophic cardiomyopathy cases in a sensitivity analysis (**Supplemental Table 3**). Interestingly, we also identified rs115740542 (P = 2.71×10^−10^) a variant near the *HFE* gene in perfect linkage disequilibrium with rs1800562 which leads to the missense change p.Cys282Tyr and is the most common cause of hereditary hemochromatosis, an iron overload disorder associated with cardiomyopathy.^31^ Additionally, rs855791 (P = 5.65×10^−10^) a variant in *TMPRSS6*, another locus involved in iron homeostasis^32^ was associated with IVS T1 time. We identified a variant in *CAMK2D* (rs55754224, P = 1.44×10^−9^) that has been previously associated with atrial fibrillation.^33^ The remaining genome-wide significant variants were located near genes associated with cardiac arrhythmias,^34–36^ cardiac remodeling^37^ and myocyte cytoskeletal proteins^38^ including *PPP2R3A, PIM1* and *KANK1*, respectively.

Four out of the 5 loci identified in the LV FW GWAS overlapped with loci in the IVS GWAS, consistent with their high genetic correlation. The top locus in the IVS GWAS, rs2627230, was borderline genome-wide significant in the LV FW GWAS (P = 8.13×10^−8^) with concordant direction of association with T1 time for both segments. The non-overlapping locus associated only with left ventricle free wall T1 time was rs369541018, an intronic variant in *KANSL1* with no known association with cardiovascular disease. This locus includes *MAPT* and has been previously described as a complex locus within one of the largest LD blocks (∼1.8 Mb) in the human genome with reported association with Alzheimer’s disease,^39^ which likely explains the high density of SNPs associated with this locus in our analysis. Re-analysis of this locus using genotype data revealed similar findings suggesting that the dense signal is not related to a local imputation error (**Supplemental Figure 14**). A quantile-quantile (QQ) plot excluding the *KANSL1* locus (**Supplemental Figure 15**) showed no early focal deviation in expected and observed p-values as was noted in the original QQ plot (**Supplemental Figure 11b**).

In conditional analysis adjusting for lead SNPs within each identified locus in the IVS and LV FW GWAS, no additional independent genome-wide significant SNPs were identified. We then performed sensitivity genome-wide association analysis of the IVS and LV FW after excluding participants with prevalent heart failure, dilated cardiomyopathy, hypertrophic cardiomyopathy or myocardial infarction. In the IVS GWAS sensitivity analysis, we found 10 genome-wide significant loci all of which overlapped with the original main IVS GWAS loci. The *KANK1* locus (rs58774558, P = 7.16×10^−8^) became borderline significant in the IVS GWAS sensitivity analysis (**Supplemental Figure 16a, 17a and Supplemental Table 3**). In the LV FW GWAS sensitivity analysis, we replicated all 5 loci in the main LV FW GWAS and identified one additional genome-wide significant locus near the *ACP1* (rs554982961, P=3.63×10^−8^) gene and a borderline significant locus near *NRP1* (rs61843241, P = 9.06 × 10^−8^) (**Supplemental Figure 16b, 17b, 18 and Supplemental Table 4**). Both *ACP1*^40^ and *NRP1*^41,42^ have been shown to play a role in cardiac hypertrophy and fibrosis. Furthermore, we performed an additional sensitivity analysis for the IVS GWAS excluding study participants with prevalent hereditary hemochromatosis (n=62) and found that the 2 loci associated with iron homeostasis, *H2BC4/HFE* and *TMPRSS6*, remained genome-wide significant (**Supplemental Figure 19, 20 and Supplemental Table 5)**.

### Transcriptome Wide Association Analysis Highlights role of *ADAMTSL1* and *SLC2A12* Expression in Myocardial Interstitial Fibrosis

Of the 14 unique lead SNPs identified in the IVS and LV FW genome wide association analyses, 9 (or their proxies, r^2^ > 0.6) were significant expression quantitative trait loci (eQTL) in the left ventricle and right atrial appendage. Implicated genes included *SLC2A12, SOD2* and *ADAMTSL1* in the left ventricle and *ADAMTSL1, CAMK2D, VEGFC* and *PPP2R3A* in the right atrial appendage (**Supplemental Table 6**). We then performed a transcriptome-wide association study (TWAS) using gene expression data from human left ventricle and right atrial appendage tissue and the IVS and LV FW GWAS summary statistics.^43,44^ TWAS results for the IVS and LV FW were concordant (TWAS results with P<0.05 are summarized in **Supplemental Tables 7 and 8**). Increased expression of *ADAMTSL1* in left ventricular tissue (P_IVS_ = 5.27×10^−9^; P_LV FW_ = 4.64×10^−8^) was associated with significant increase in myocardial interstitial fibrosis as measured by native myocardial T1 time. On the other hand, increased expression of *SLC2A12* in the left ventricle (P_IVS_ = 1.89×10^−10^; P_LV FW_ = 6.59×10^−7^) was associated with significant decrease in myocardial interstitial fibrosis (**Figure 4c and 4d**). In the right atrium, increased expression of *ADAMTSL1* was associated with significant increase in myocardial interstitial fibrosis (P_IVS_ = 1.29×10^− 7^; P_LV FW_ = 3.77×10^−8^) (**Supplemental Table 8**).

## DISCUSSION

We developed an automated machine-learning model to measure myocardial interstitial fibrosis in over 42,000 participants in the UK Biobank. We identified associations between myocardial fibrosis and impaired glucose metabolism, systemic inflammation, renal disease, aortic stenosis, cardiomyopathy, atrial fibrillation, and conduction disease. Furthermore, greater myocardial fibrosis was an independent predictor of incident atrial fibrillation, heart failure and conduction disease. In the first large-scale genome-wide association study of native myocardial T1 time in the human heart, we identified 12 independent loci implicating genes involved in biological pathways relevant to fibrosis including glucose homeostasis (*SLC2A12*), iron homeostasis (*HFE, TMPRSS6*), tissue repair (*ADAMTSL1, VEGFC*), oxidative stress (*SOD2*), cardiac hypertrophy (*MYH7B*) and, calcium signaling (*CAMK2D*). Overall, the heritability of myocardial interstitial fibrosis as measured by native myocardial T1 time was relatively low, emphasizing the important contribution of non-genetic environmental factors to cardiac fibrosis. Our findings prioritize a number of pathways relevant to myocardial fibrosis for further investigation.

Our findings have several major implications. First, our results highlight the salient role of glucose homeostasis and diabetes in propagation of myocardial fibrosis. Our strongest GWAS locus *SLC2A12* encodes a highly expressed insulin-independent glucose transporter, GLUT12, in the heart,^22^ and diabetes mellitus types 1 and 2 were among the diseases most strongly associated with increased myocardial T1 time in this study. GLUT12 knock-out in zebrafish leads to development of heart failure and a diabetic phenotype,^24^ consistent with our TWAS results suggesting that decreased expression of *SLC2A12* in cardiac tissue was associated with increased interstitial fibrosis. The role of hyperglycemia in the pathogenesis of diabetic cardiomyopathy via accumulation of advanced glycation end-products (AGE) that cross-link extracellular matrix proteins and transduce profibrotic signals through activation of receptor for AGE-mediated pathways is well-established.^45^ Additionally, the salutary effects of anti-diabetic therapies that block sodium-glucose cotransporter-2 on outcomes of heart failure with both reduced^46^ and preserved^47^ ejection fraction, irrespective of concomitant diabetes, have been recently recognized. Our findings further highlight the role of glucose homeostasis in myocardial fibrosis in the human heart and pinpoint potential additional pathways that warrant further interrogation.

Second, pathways involved in tissue repair including extracellular matrix turnover and lymphangiogenesis were associated with myocardial fibrosis. Increased human cardiac expression of *ADAMTSL1*, an ADAMTS-like protein which lacks catalytic activity and is thought to modulate the function of ADAMTS metalloproteinases with integral roles in extracellular matrix turnover,^28,29^ was associated with higher myocardial interstitial fibrosis in this study. The exact effect of ADAMTSL-1 on ADAMTS metalloproteinases remains unknown; however, homology between mammalian ADAMTSL-1 and invertebrate papilin, a known inhibitor of ADAMTS-2, has been reported.^48,49^ Studies in mice with cardiac-specific overexpression of *Adamts2* have shown an abrogated pressure overload-induced hypertrophic response.^50^ Thus, potential inhibition of ADAMTS-2 activity may explain the increased myocardial fibrosis associated with increased expression of ADAMTSL-1 in the human myocardium. *VEGFC-* and *VEGFD*-mediated lymphangiogenesis has been associated with cardiac repair and knock-out zebrafish models go on to develop severe cardiac hypertrophy and myocardial interstitial fibrosis. Additionally, use of systemic VEGF inhibitor therapy, such as bevazicumab, has been associated with a 2-4% risk of incident heart failure.^51^ In this study, the lead SNP rs365843, tagging *VEGFC* was associated with increased expression of *VEGFC* in human right atrial appendage tissue and with lower myocardial native T1 times reflecting lower myocardial interstitial fibrosis. Thus, our current findings extend those from animal models and suggest a role for reparative pathways involving *ADAMTS* and *VEGFC* in reducing myocardial fibrosis in the human heart.

Third, our results shed light on the role of myocardial oxidative stress and inflammation in development of myocardial interstitial fibrosis. rs9457699, a lead SNP in the IVS native myocardial T1 time GWAS, is an eQTL for *SOD2* in the human left ventricle and was associated with increased expression of *SOD2* and lower native myocardial T1 time. This is congruent with findings from *Sod2* knockout mice which exhibit increased levels of oxygen reactive species with associated myocardial fibrosis and development of dilated cardiomyopathy.^26^ Furthermore, we found an association between rheumatoid arthritis as well as baseline C-reactive protein (a well-established clinical inflammatory marker) with increased native myocardial T1 time emphasizing the role of inflammation in the propagation of myocardial interstitial fibrosis.

Fourth, a number of additional established pathways involved in myocardial fibrosis were implicated in our results. We provide further evidence for the role of *CAMK2D* in pathologic cardiac remodeling and fibrosis^52,53^ via association of *CAMK2D* with increased myocardial T1 time. Additionally *PIM1*,^54^ *NRP-1*,^41,42^ and *ACP1*^40^ have been shown to play a role in myocardial fibrosis and were implicated in our study results.

Notably, we identified two genes, *HFE* and *TMPRSS6*, associated with iron homeostasis in our analysis of the genetic determinants of myocardial T1 time. These loci remained significantly associated with myocardial T1 time after excluding individuals with prevalent hereditary hemochromatosis. Cardiac T1 mapping is a known sensitive marker of iron deposition in the heart that complements T2* especially in early stages of iron overload which could be missed by T2*.^55^ Iron deposition in the heart alters myocardial tissue magnetic properties and is associated with lower T1 time and with development of iron overload cardiomyopathy.

Despite the inherent noise associated with native myocardial T1 time that may have limited findings from prior studies, we found that examining myocardial native T1 time at scale yielded biologically plausible insights into validated pathways involved in myocardial fibrosis. The laborious manual segmentation required for T1 time measurement has limited its availability mostly to small research-based cohorts.^10^ We demonstrate the feasibility of automated quantification of myocardial T1 time at scale which promises to accelerate our understanding of this relatively under-studied phenotype. Additionally, we provide evidence that myocardial T1 mapping identifies myocardial changes associated with a number of systemic and cardiovascular diseases beyond standard measures of LV size and function and is of prognostic value in predicting risk of incident cardiovascular disease. These findings bolster the diagnostic and prognostic value of T1 mapping and advocate for more widespread use and study of myocardial T1 mapping.

Our study has several limitations. First, the cardiac MRI protocol in the UK Biobank did not include the use of contrast agents which prohibited calculation of extracellular volume fraction. In some studies, extracellular volume fraction has been shown to be a more sensitive marker of myocardial interstitial fibrosis and have stronger association with disease as compared to native T1 time.^56^ Second, the T1 maps are obtained at a single mid-ventricular short-axis slice and, while T1 mapping aims to measure diffuse non-focal fibrosis, we cannot be certain that a single slice is representative of myocardial fibrosis throughout the left ventricle. Third, while increased native myocardial T1 time usually reflects increases in extracellular volume due to fibrous tissue deposition, it can also be elevated in the presence of tissue edema. Fourth, the UK Biobank study population is predominantly of European ancestry and findings from our genetic analysis may not necessarily apply to other ancestries. Fifth, longer follow-up and continued imaging of UK Biobank participants will allow for more powered analyses examining the prognostic role of myocardial interstitial fibrosis in predicting incident cardiovascular disease than was possible in the current study.

In conclusion, machine learning enables quantification of myocardial interstitial fibrosis at scale.

Our study yields insights into novel biological pathways underlying cardiac fibrosis and prioritizes a number of pathways relevant to myocardial fibrosis for further investigation.

## ONLINE METHODS

In the sections below, we provide a detailed description of the methods used in this manuscript. Briefly, we trained a machine learning model to segment cardiac T1 maps from the UK Biobank and measure T1 time at the IVS and LV FW. We examined the associations between native myocardial T1 time and cardiometabolic risk factors and cardiovascular disease. We then performed a genome and transcriptome-wide association analysis of native myocardial T1 time at the IVS and LV FW.

### Study Design and Population

The UK Biobank is a prospective cohort of 502,629 individuals from the UK enrolled between 2006-2010 with deep phenotyping, imaging and multiple genomic data types. The cohort design has been previously described.^15,57^ Briefly, around 9.2 million individuals 40-69 years old living in England, Scotland, and Wales were invited to participate in the study and 5.4% agreed to participate. Extensive questionnaire data, physical measures, and biological samples were collected at baseline, with ongoing data collection in large subsets of the cohort, including repeated assessments and multimodal imaging. Starting in 2014, 42,654 participants have returned for the first multi-modal imaging visit including cardiac magnetic resonance imaging with T1 mapping allowing for the assessment of myocardial interstitial fibrosis.^14^ All study participants are followed longitudinally for health-related outcomes through linkage to national health-related datasets.

Use of UK Biobank data was performed under application number 17488 and was approved by the local Massachusetts General Hospital institutional review board.

### Cardiovascular Magnetic Resonance Imaging T1 Mapping Protocol and Image Quality Control

A standardized non-contrast enhanced cardiac magnetic resonance imaging protocol using a clinical wide bore 1.5 Tesla scanner (MAGNETOM Aera, Syngo Platform VD13A, Siemens Healthcare, Erlangen, Germany) is performed on all cardiac MRI substudy participants. The scanner is equipped with 48 receiver channels, a 45 mT/m and 200 T/m/s gradient system, an 18 channel anterior body surface coil used in combination with 12 elements of an integrated 32 element spine coil and electrocardiogram gating for cardiac synchronization. The imaging protocol includes: 3 long-axis cines, 1 short-axis cine, phase contrast sequence at the left ventricular outflow tract, 3 segment short-axis tagging and midventricular short-axis T1 mapping. Native T1 mapping within a single breath hold was performed using the Shortened Modified Look-Locker Inversion recovery (ShMOLLI, WIP780B) technique. The following imaging parameters for T1 mapping were implemented: field of view 360 × 236 mm, voxel size 0.9 × 0.9 × 8.0, flip angle 35 degrees and TR/TE 2.6/1.07 ms.^14^ T1 maps were generated online and stored in the UK Biobank imaging database.

To date, the UK Biobank MRI core lab has only released raw T1 maps to UK Biobank researchers. As such, we developed our own automated pipeline to measure native myocardial T1 time from raw T1 maps (**Figure 1**). First, we set up an automatic procedure to identify raw T1 map series among the several files provided by the UK Biobank under the category “Experimental shMOLLI sequence images” (UK Biobank Field ID 20214). Preliminary explorations indicated that the T1 map series names contained the “t1map” keyword and explicitly mentioned the MRI “sax (short-axis)” view. Therefore, we discarded all series with names not containing either of the two keywords. Then, we set up a standardized quality control process using a custom online tool to streamline the review of the 42,654 selected T1 maps by four experienced MRI reviewers (V.N, M.D.R.K, P.D.A., and J.W.C., **Supplemental Figure 21**). Two myocardial segments of interest were defined for this analysis, the IVS and the LV FW, to allow for examination of regional variation in myocardial interstitial fibrosis and region-specific association with disease. Additionally, this enabled the characterization of the genetic determinants of septal vs. free wall myocardial interstitial fibrosis.

All images were reviewed and assessed for overall image quality as well as artifacts involving segments of interest (IVS and LV FW). A cardiologist (V.N.) reviewed all images that were flagged by any of the four reviewers and made a final ascertainment on image quality and extent of artifact involving segments of interest. Off-axis images and those with severe distortion of overall image pixel intensity were excluded (**Supplemental Figure 22**). Artifact within a segment of interest was deemed major if it affected at least one-third of the segment of interest (**Supplemental Figure 22)**. Segments of interest with major artifacts were excluded from the reported T1 times for that segment of interest. Of 42,654 individuals who underwent cardiac T1 mapping, 1,149 and 3,343 had major artifacts involving the interventricular septum or left ventricle free wall, respectively, and were excluded from the analysis involving the affected segment (**Supplemental Figure 1**). We found that the LV FW segment had a 3-fold higher incidence of artifact as compared to the interventricular septum. The artifact-prone nature of the LV FW segment is well recognized in the field of cardiac T1 mapping and has led some experts to advocate for preferential use of septal T1 time to differentiate health and disease states of the myocardium.^16,17^ As such, while we present results for both the IVS and LV FW, the IVS native myocardial T1 time constituted our primary analysis.

### Semantic Segmentation, Region of Interest Selection and Native Myocardial T1 Time Measurement

Six hundred (500 training, 100 validation) T1 maps were randomly selected and used to develop our machine learning model. Two cardiologists, V.N. and J.W.C., labeled all cardiac structures within the short-axis T1 maps (350 V.N.; 250 J.W.C.). Fifty T1 maps were labeled by both readers to allow for assessment of inter-reader reliability. Cardiac structures that were labeled included: interventricular septum, LV free wall, papillary muscles, LV blood pool, right ventricle free wall and right ventricle blood pool. Additionally, a region of interest encompassing the mid-myocardium across the segment of interest and excluding the endocardial and epicardial borders was delimited separately within the IVS and LV FW (**Supplemental Figure 2**). Pixel intensity values were transformed to T1 times using the accompanying T1 map legend. IVS and LV FW native myocardial T1 time was measured as the median T1 time for all pixels within the corresponding region of interest.

### Machine Learning Model Development

For segmenting cardiac structures in cardiac MRI T1 maps, we employed the DenseNet-121 architecture^58^ as the base encoder model in a U-Net model^59^ that was pre-trained on ImageNet.^60^ DenseNets are constructed with two principal building blocks: (1) dense blocks comprising of batch normalization, the non-linear ReLU activation function, and 3×3 convolutions of increasing number of channels that are propagated from previous layers to enable efficient gradient flow; and (2) transition blocks that compress the number of channels by half using channel-wise convolutions (1×1), and perform a spatial reduction by a factor of 2 by using an average pooling layer of stride 2 and pool size 2. The U-Net architecture contains long-range skip connections that allow for pixel-accurate segmentation by sharing feature information along a contracting-expansive path. This is achieved by concatenating features at each downsampling in the encoder with the corresponding features at each upsampling step. These ‘skip connections’ preserve contextual and spatial information.

The inputs for this model were the cardiac MRI T1 maps with size 288 × 384 × 3. The models were trained with the Adam optimizer^61^ with a learning rate set to a cosine decay policy decaying from 0.0001 to 0 over 100 epochs, weight decay of 0.0001, categorical cross-entropy as the loss function, and a batch size of 16. No additional hyperparameter search or ablation studies were performed.

For all training data, the following augmentations (random permutations of the training images) were applied: random shifts in the XY-plane by up to ±16 pixels and rotations by up to ±5 degrees around its center axis.

### Phenotypic Characterization of the Study Sample and Association with Native T1 Time

Prevalent cardiometabolic, cardiovascular and systemic inflammatory diseases at time of first visit for cardiac MRI as well as incident cardiovascular events were ascertained using International Classification of Diseases, 9th and 10th editions, codes and Office of Population Censuses and Surveys (OPCS) Classification of Interventions and Procedures version 4 codes (**Supplemental Table 9**). Derived myocardial T1 times were rank-based inverse normal transformed. Multiple linear regression was used to assess the association of prevalent cardiometabolic, systemic inflammatory and cardiovascular disease at time of MRI with native myocardial T1 time adjusting for age at MRI visit, sex, height, weight, beta blocker therapy, and angiotensin-converting enzyme inhibitor/angiotensin receptor blocker therapy. We additionally performed a sensitivity analysis with incremental adjustment for LV mass and LV ejection fraction to examine whether the association between T1 time and examined diseases was independent of standard measures of left ventricular structure and function.

A time-to-event analysis was performed to assess the association of native myocardial T1 time with incident cardiovascular events. Follow-up time was defined as time from MRI visit to first occurrence of the outcome of interest, death or last follow-up (April 30th, 2020). For each incident disease analysis, study participants with prevalent disease at time of MRI were excluded as they were not at risk for the outcome of interest. We then stratified the cohort into the upper 20th and lower 80th percentile of native myocardial T1 time. Using a multivariable Cox proportional hazards model adjusted for age at MRI, sex, height, weight, beta blocker therapy, and angiotensin-converting enzyme inhibitor/angiotensin receptor blocker therapy, we examined the association of native myocardial T1 time with incident cardiovascular events. Adjusted Kaplan-Meier curves were constructed to compare incidence rate of cardiovascular events between the two groups. The validity of the proportional hazards assumption was verified by examining the Schoenfeld residuals. All statistical tests were performed using R version 4.0.2 (R Foundation for Statistical Computing, Vienna, Austria)(R, Core Team 2020) and two-sided P-values <0.05 were considered statistically significant.

### Genotype data, Imputation, Sample and Variant Quality Control

In total, 488,377 UK Biobank participants were genotyped using either one of two overlapping arrays, the UK BiLEVE Axiom Array or the UK Biobank Axiom Array. Prior to imputation, a number of quality control filters were applied to the genotype data. Variants with >5% missing rate, minor allele frequency <0.0001 and that violated Hardy-Weinberg Equilibrium (P-value threshold <1×10^−12^) were excluded. Additionally, samples that were identified as outliers for genotype missingness rate (>5%) and heterozygosity were also excluded. These filters resulted in a genotype dataset that included 670,730 autosomal variants in 487,442 samples. Imputation into the Haplotype Reference Consortium (HRC) and UK10K+ 1000G phase 3 reference panels was carried out using IMPUTE4. The imputation process resulted in a dataset with 93,095,623 autosomal SNPs and short indels in 487,442 individuals.^15^

Of 42,654 study participants who underwent cardiac magnetic resonance imaging with T1 mapping, 41,635 had imputed genetic data available. Sample and variant quality control filters were applied prior to conducting genetic association analyses. Samples with sex chromosome aneuploidy and those with discordant genetically inferred and self-reported sex were excluded. One of each pair of third degree relatives or closer was excluded. Variants with imputation quality score (INFO) <0.3 and those with minor allele frequency <0.1 were excluded. Following quality control, our dataset included 40,399 individuals with 9,853,972 SNPs and short indels. Among the 40,399 study participants with adequate quality genetic data, 39,339 and 37,306 had IVS and LV FW native myocardial T1 time data that passed quality control available, respectively (**Supplemental Figure 1**).

### Genome-wide Common Variant Association Analysis Methods

We performed a common variant genome wide association analysis of both interventricular septum and LV free wall native myocardial T1 time using a fixed effect linear regression model in PLINK 2.0.^62^ The models were adjusted for age at MRI, sex, the unique MRI scanner serial number, genotyping array, and first ten principal components of genetic ancestry. Rank-based inverse normal transformation was applied to the measured myocardial T1 times from each myocardial segment (IVS and LV FW). As such, effect size estimates in the GWAS are dimensionless and reflect approximately multiples of 1 standard deviation of the underlying quantitative trait. A two-sided P-value <5×10^−8^ was used to define genome-wide significant common variants and 5×10^−8^ < P-value < 1×10^−6^ denoted suggestive loci. Distinct genomic loci were defined by starting with the SNP with the lowest p-value, excluding other SNPs within 500 kb, and iterating until no SNPs remained. The independently significant SNPs with the lowest p-value at each genomic locus are termed lead SNPs. We then performed a conditional analysis adjusting for the imputed allele dosage of each lead SNP to examine for additional independent genome-wide significant SNPs within a locus. We performed two sensitivity analyses. First, we repeated the above GWAS of IVS and LV FW myocardial T1 time after exclusion of individuals with prevalent heart failure, dilated cardiomyopathy, hypertrophic cardiomyopathy or myocardial infarction at time of MRI (N_IVS GWAS_=38,339, N_LV FW GWAS_=36,381). Second, we repeated the IVS GWAS after excluding participants with prevalent hereditary hemochromatosis (n=62) to examine whether the identified genome-wide significant loci associated with iron homeostasis were driven by hereditary hemochromatosis cases.

Linkage disequilibrium (LD) score regression analysis was performed using ldsc version 1.0.0.^63^ With ldsc, the genomic control factor (lambda GC) was partitioned into components reflecting polygenicity and inflation, using the software’s defaults.

Regional association plots were generated with LocusZoom^64^ using LD data from the 1000G phase 3 European reference panel. In instances where lead SNPs were not part of the 1000G phase 3 reference panel, in-sample LD was calculated using PLINK 1.9.^62^

### Heritability and Genetic Correlation Analysis

SNP-heritability of the IVS and LV FW native myocardial T1 time was assessed using BOLT-REML v2.3.4.^65^ We also computed genetic correlation between native myocardial T1 time for each segment and other MRI parameters including LV end diastolic and systolic volumes, LV mass, LV ejection fraction, left atrial end diastolic and systolic volumes and left atrial ejection fraction using ldsc version 1.0.0.^66^ These cMRI-based phenotypes from the UK Biobank have been described previously.^21,67,68^

### Expression Quantitative Trait Locus and Transcriptome-Wide Association Analysis Methods

We performed an expression quantitative trait locus look-up using version 8 of the Genotype-Tissue Expression (GTEx) database.^43^ In-sample LD was calculated for all variants within 1 MB of genome-wide significant lead SNPs using PLINK 1.9. List of proxy SNPs for each lead SNP were generated using an LD r^2^ threshold of >0.6. We searched the GTEx v8 database for statistically significant differential gene expression in right atrial appendage and left ventricular tissues associated with the lead SNPs and their proxies. When no significant differential gene expression associated with the lead SNP was identified, significant findings from the closest proxy were reported.

We then performed a transcriptome-wide association analysis to test the mediating effects of gene expression levels in right atrial appendage and left ventricular tissue on native myocardial T1 time. We used pre-computed transcript expression reference weights derived using elastic net models from S-PrediXcan on GTEx v8 eQTL data for the right atrial appendage and left ventricle.^44^ S-Predixcan was then run with its default settings. A Bonferroni-corrected P-value threshold <7.5×10^−6^ (0.05/6,637 genes tested) was used to define significant gene expression-phenotype associations.

## Data Availability

UK Biobank data are made available to researchers from research institutions with genuine research inquiries, following IRB and UK Biobank approval. All other data are contained within the article and its supplementary information, or are available upon reasonable request to the corresponding author.

## Code Availability

Code used to ingest, quality control, and train machine learning models are available at https://github.com/broadinstitute/ml4h under an open-source BSD license.

## Author Contributions

V.N., J.W.C., P.T.E. and S.A.L. conceived the study. M.D.R.K. and P.D.A. ingested and prepared cardiac MRI data. V.N., M.D.R.K., P.D.A., and J.W.C., performed quality control. M.D.R.K. trained machine learning models. V.N., M.D.R.K., and P.D.A., performed the main analyses. V.N., M.D.R.K., P.D.A., J.W.C., P.T.E. and S.A.L. wrote the paper. All other authors contributed to the analysis plan or provided critical revisions.

