## Supplemental Figures for "Genetics of Myocardial Interstitial Fibrosis in the Human Heart and Association with Disease"

### Supplemental Figure Legends

**Supplemental Figure 1.** Study flowchart. GWAS: genome wide association study; IVS: interventricular septum; LV FW: left ventricle free wall; QC: quality control.

**Supplemental Figure 2.** Manually-traced and labelled cardiac structures within mid-ventricular short-axis T1 maps. IVS: interventricular septum; LV: left ventricle; ROI: region of interest; RV: right ventricle.

**Supplemental Figure 3.** Inter-reader correlation of manually-derived T1 time of the interventricular septum (a) and left ventricle free wall (b) in 50 overlapping T1 maps.

**Supplemental Figure 4.** Correlation of manually- and deep learning model-derived T1 time of the interventricular septum (a) and left ventricle free wall (b) in the validation set (n=100).

**Supplemental Figure 5.** Sex-specific native myocardial T1 time distribution of the interventricular septum (a) and left ventricle free wall (b).

**Supplemental Figure 6.** Correlation of left ventricle free wall and interventricular septum T1 time among 39,113 participants with available T1 time data for both segments. LV FW: left ventricle free wall; IVS: interventricular septum.

**Supplemental Figure 7.** Forest plot depicting multivariable adjusted change in native myocardial T1 time associated with prevalent cardiovascular, metabolic and systemic inflammatory diseases at time of cardiac MRI compared to healthy controls. Numbers in parentheses represent number of cases for each disease included in the analysis examining association with the interventricular septum and left ventricle free wall T1 time, respectively. 27,291 and 25,881 healthy controls with interventricular septum and left ventricle free wall T1 time data available, respectively, constituted the reference groups. 38,731/36,663 had data on both C-reactive protein and interventricular septum/left ventricle free wall T1 time and were included in this analysis.

**Supplemental Figure 8.** Forest plot depicting multivariable adjusted change in native myocardial T1 time associated with prevalent cardiovascular, metabolic and systemic inflammatory diseases at time of cardiac MRI compared to healthy controls after incremental adjustment for left ventricular mass and ejection fraction in the subgroup of participants who had this data available ($N_{IVS}$ = 40,231 ; $N_{LV\ FW}$ = 38,127). Numbers in parentheses represent number of cases for each disease included in the analysis examining the association with the interventricular septum and left ventricle free wall T1 time, respectively. 26,510 and 25,153 healthy controls with interventricular septum and left ventricle free wall T1 time data available, respectively, constituted the reference groups. 37,555/35,571 had data on both C-reactive protein and interventricular septum/left ventricle free wall T1 time and were included in this analysis.

**Supplemental Figure 9.** Multivariable adjusted association of native myocardial T1 time with incident cardiovascular disease. Comparisons for both interventricular septum and left ventricle free wall T1 time are between the top 20th percentile and lower 80th percentile of the T1 time distribution in the study sample. Numbers in parentheses represent number of cases for each disease included in the analysis examining the association with the interventricular septum and left ventricle free wall T1 time, respectively.

**Supplemental Figure 10.** Genetic correlation of cardiac MRI traits. Genetic correlation was calculated using LD score regression.

**Supplemental Figure 11.** Quantile-quantile plot for interventricular septum (a) and left ventricle free wall (b) native T1 time GWAS.

**Supplemental Figure 12.** Regional association plots of genome-wide significant loci from the interventricular septum T1 time GWAS.

**Supplemental Figure 13.** Regional association plots of genome-wide significant loci from the left ventricle free wall T1 time GWAS.

**Supplemental Figure 14.** *KANSL1* locus regional association plot using genotyped (a) and imputed (b) genomic data. (a) The lead SNP for the genetic association analysis using genotyped data is rs56192752 and is used as the reference SNP for the regional association plot using genotyped data. (b) The lead SNP for the genetic association analysis using imputed data is rs369541018 (not present in genotyped dataset) and is used as the reference SNP for the regional association plot using imputed data. The lead SNP from the genotype analysis, rs56192752, is labeled in this plot to facilitate comparison of the two regional association plots.

**Supplemental Figure 15.** Quantile-quantile plot for left ventricle free wall native T1 time GWAS excluding *KANSL1* locus.

**Supplemental Figure 16.** Interventricular septum (a) and left ventricle free wall (b) native T1 time genome-wide association results across 22 autosomes after excluding participants with prevalent myocardial infarction, heart failure or dilated/hypertrophic cardiomyopathy. Nearest genes are used for annotation. The dashed grey line represents the threshold for genome-wide significance ($P<5x10^{-8}$). Number of study participants included in interventricular septum GWAS was 38,339 and in left ventricle free wall GWAS 36,381.

**Supplemental Figure 17.** Quantile-quantile plot for interventricular septum (a) and left ventricle free wall (b) native T1 time GWAS following exclusion of participants with prevalent myocardial infarction, heart failure or dilated/hypertrophic cardiomyopathy.

**Supplemental Figure 18.** Regional association plots of the two additional loci (*ACP1* and *NRP1*) identified in the left ventricle free wall genome wide association sensitivity analysis.

**Supplemental Figure 19.** Interventricular septum native T1 time genome-wide association results across 22 autosomes after excluding participants with prevalent hereditary hemochromatosis. Nearest genes are used for annotation. The dashed grey line represents the threshold for genome-wide significance ($P<5x10^{-8}$). Number of study participants included in the interventricular septum GWAS was 39,277.

**Supplemental Figure 20.** Quantile-quantile plot for interventricular septum native T1 time GWAS following exclusion of study participants with prevalent hemochromatosis.

**Supplemental Figure 21.** Screenshot of the custom online interface used to collect the quality control annotations from the MRI reviewers. The interface presents the annotators with the raw T1 map reconstruction. An annotator records the presence of local major artifacts at the left ventricle free wall or at the interventricular septum, as well as more global artifacts such as off-axis images and unphysiologically low native T1 times.

**Supplemental Figure 22.** Examples of local and global T1 map artifacts identified during the cardiac magnetic resonance imaging QC step. *Top left,* major artifact involving the interventricular septum. *Top middle,* major artifact involving the entire left ventricle free wall. *Bottom left,* the MRI acquisition was flagged as “off-axis”, being at the base of the heart and including the left ventricle outflow tract. *Bottom middle,* the T1 map reconstruction artifact yields unphysiologically low T1 times (i.e., a “dark” image) for the entire MRI slice. *Bottom right,* the entirety of the T1 map is compromised by several artifacts.

Supplemental Figure 1.

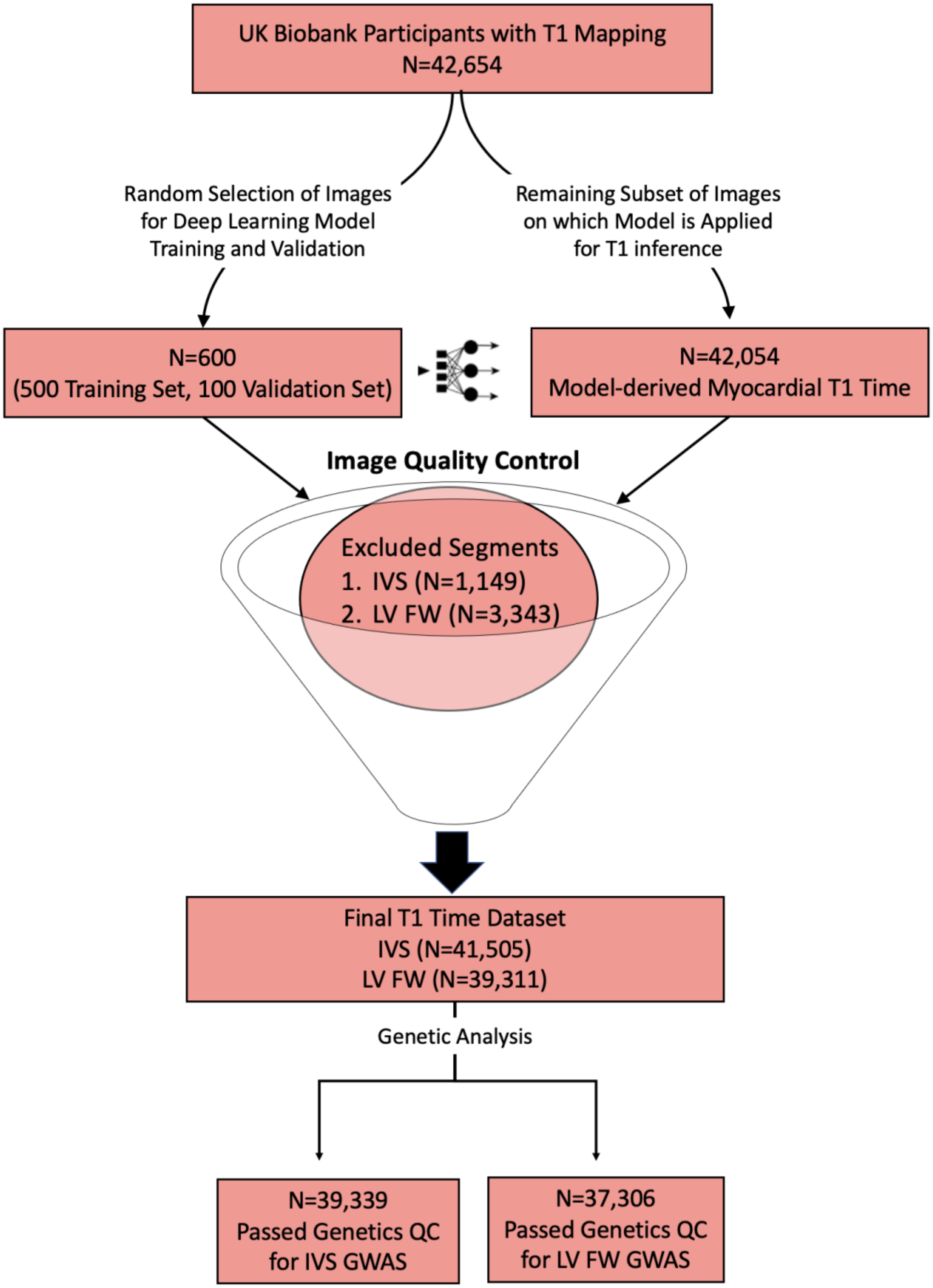

Supplemental Figure 2.

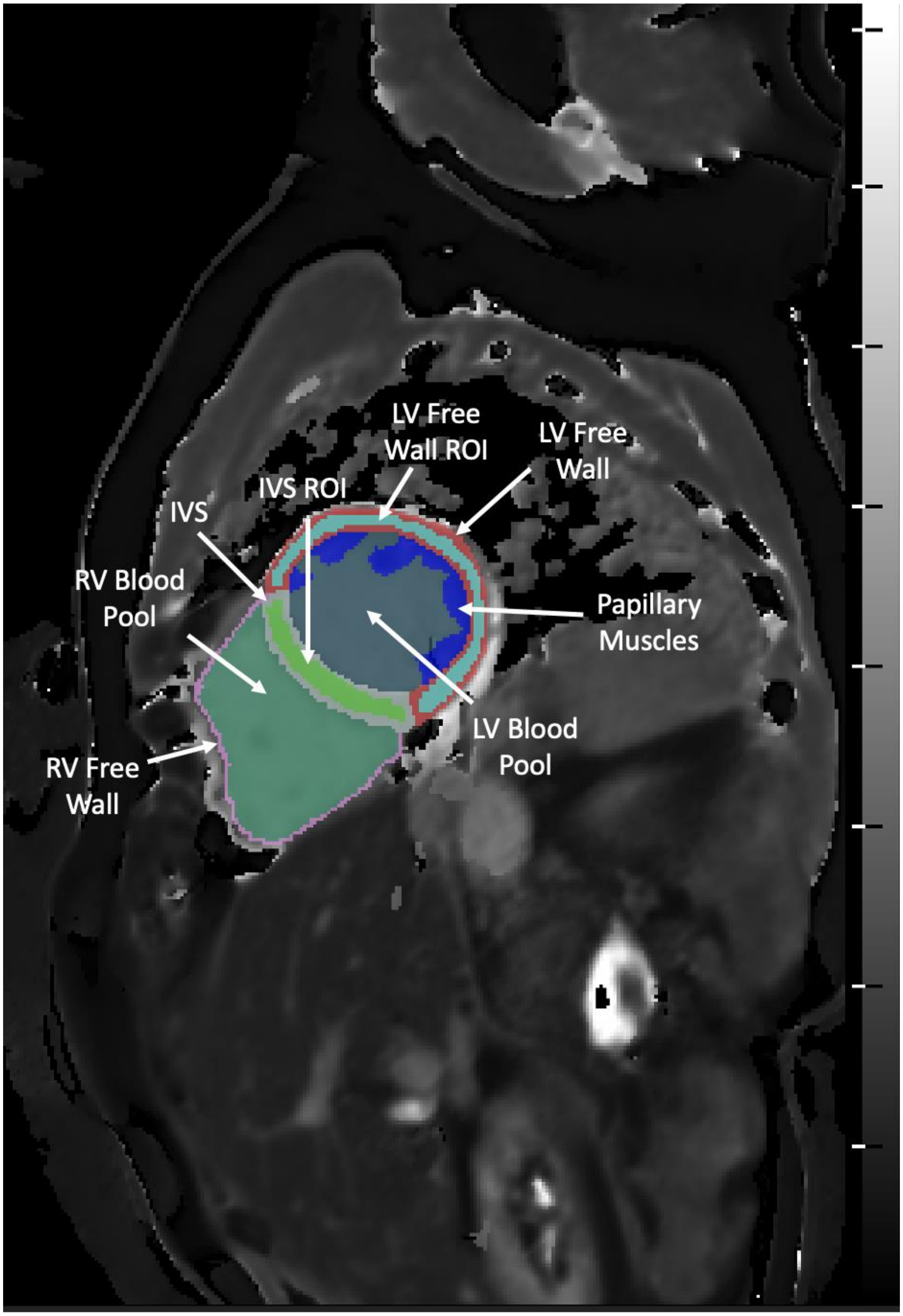

Supplemental Figure 3.

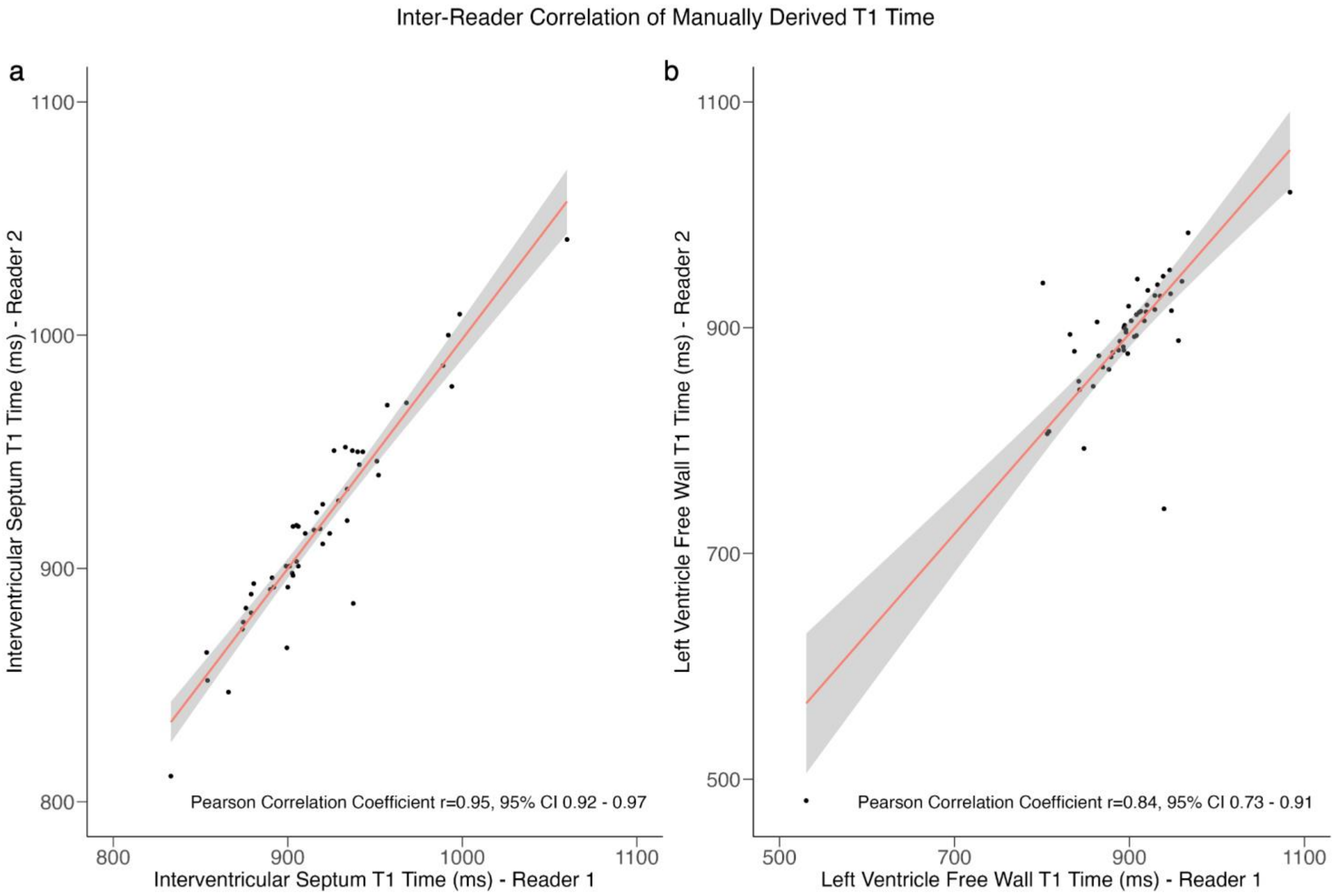

Supplemental Figure 4.

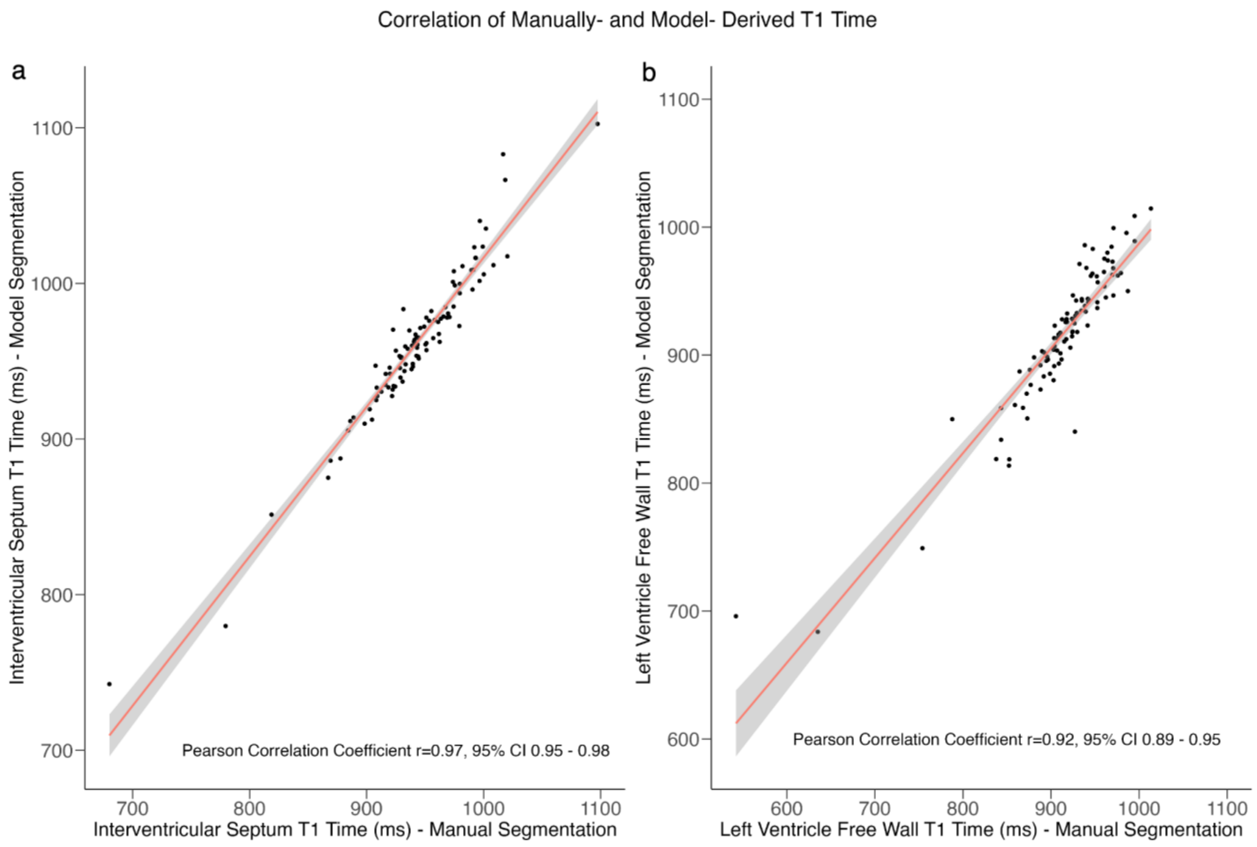

Supplemental Figure 5.

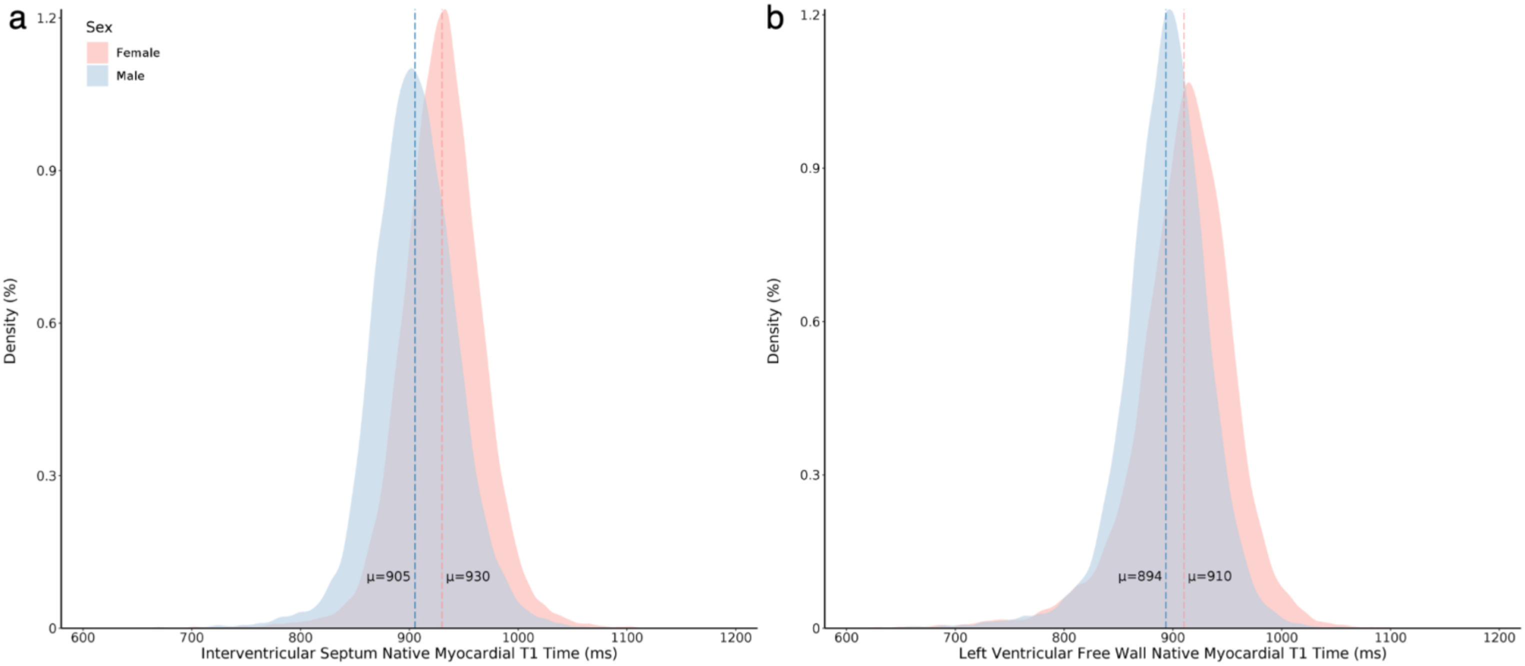

Supplemental Figure 6.

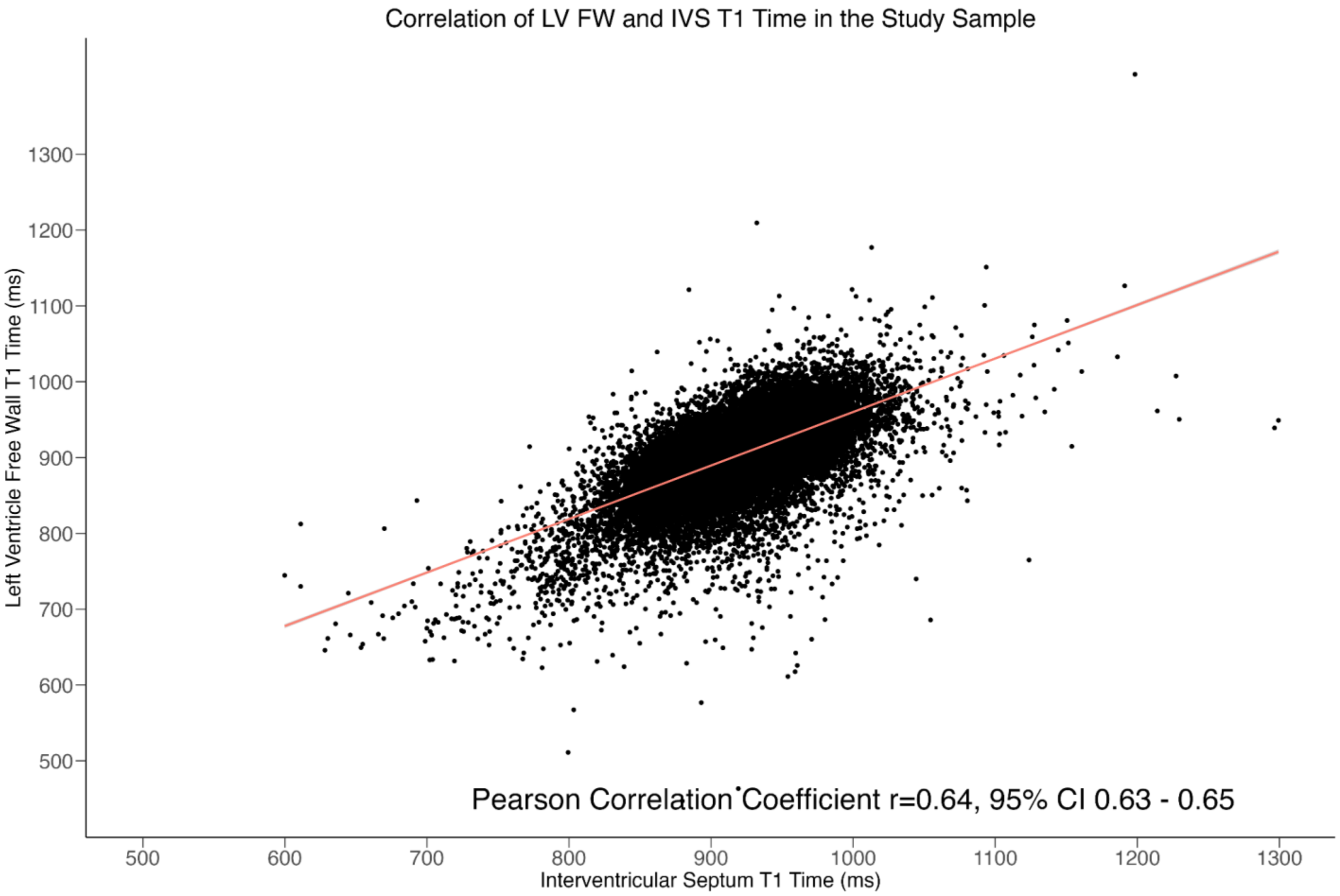

Supplemental Figure 7.

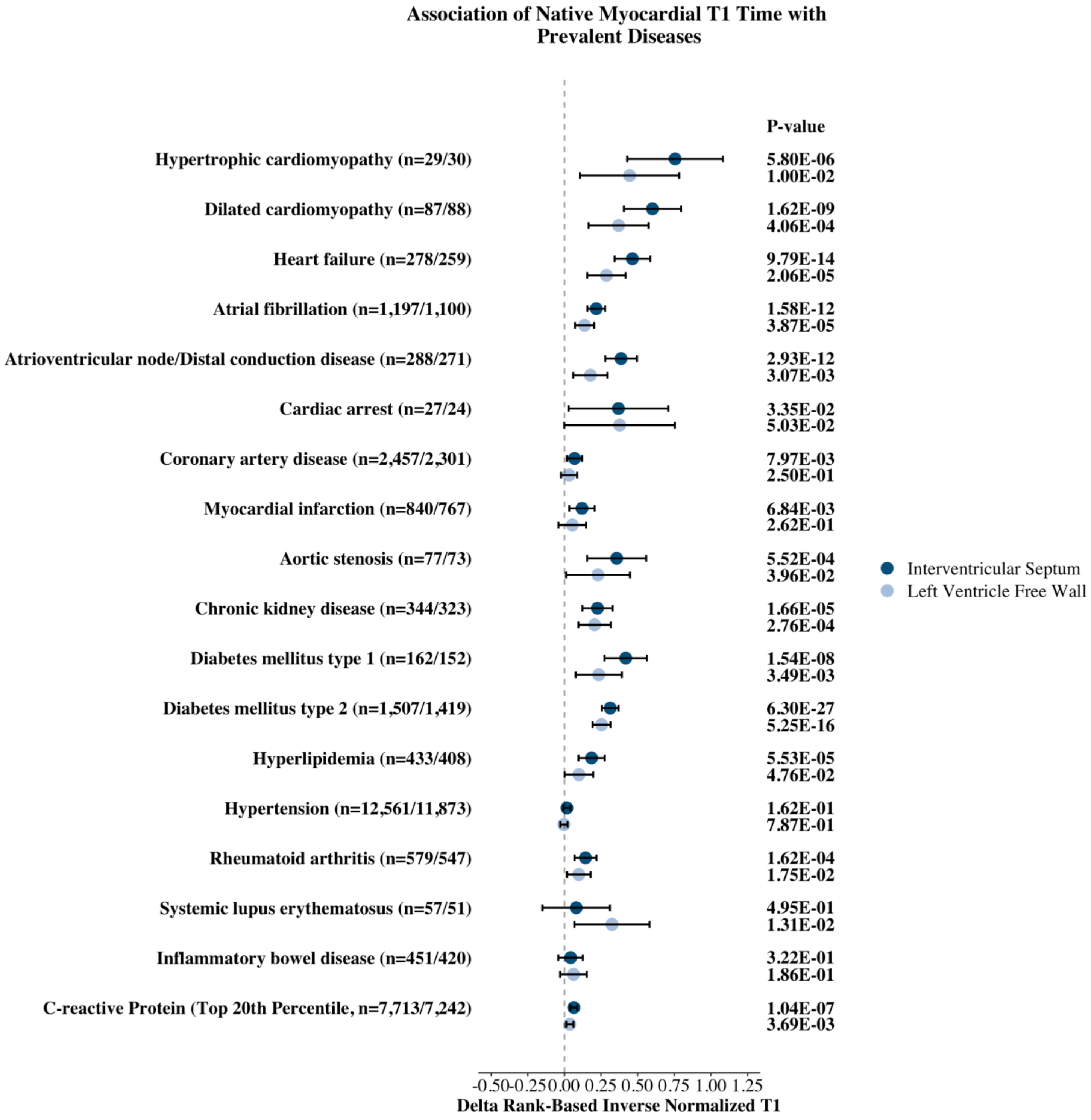

Supplemental Figure 8.

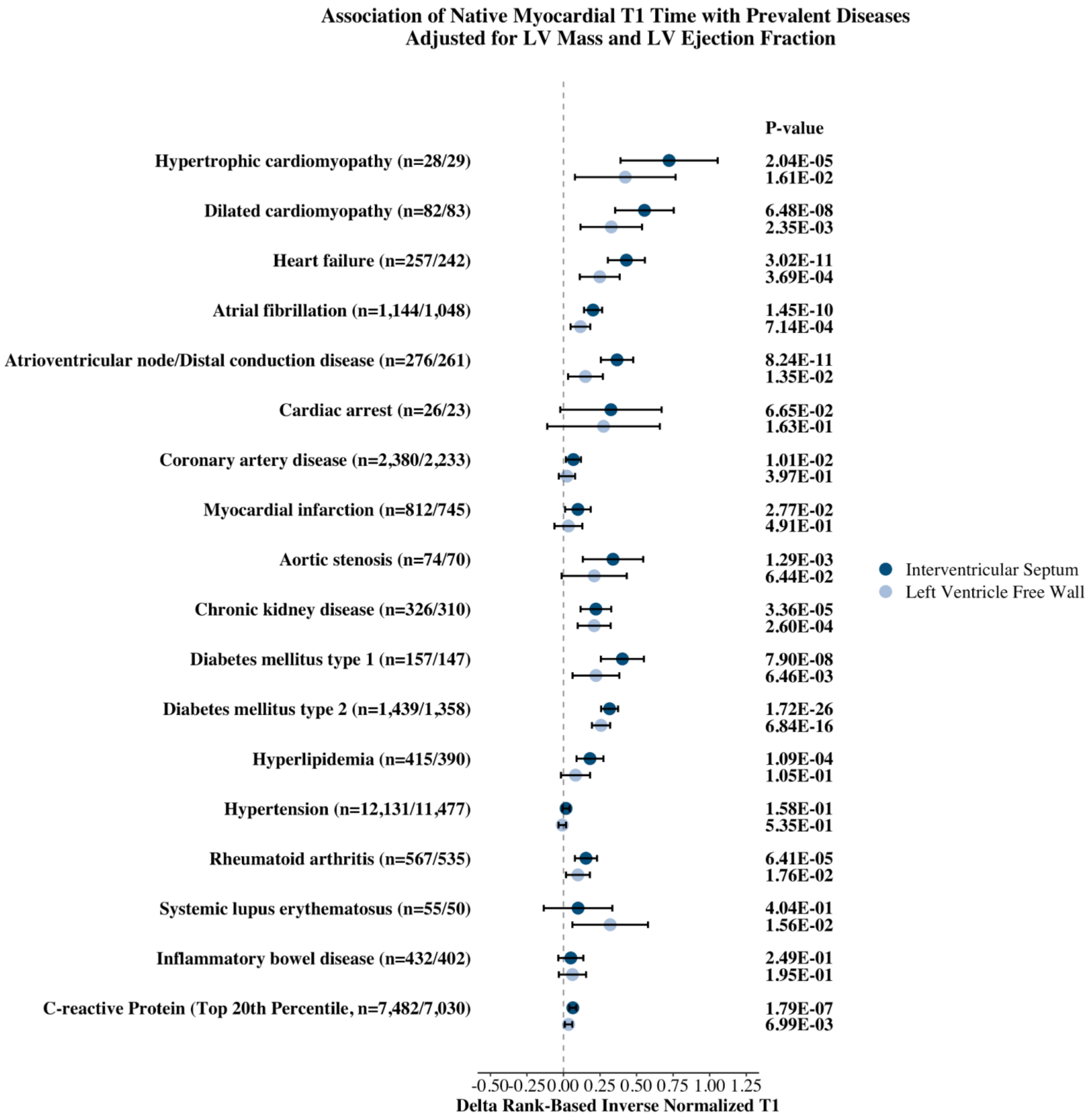

Supplemental Figure 9.

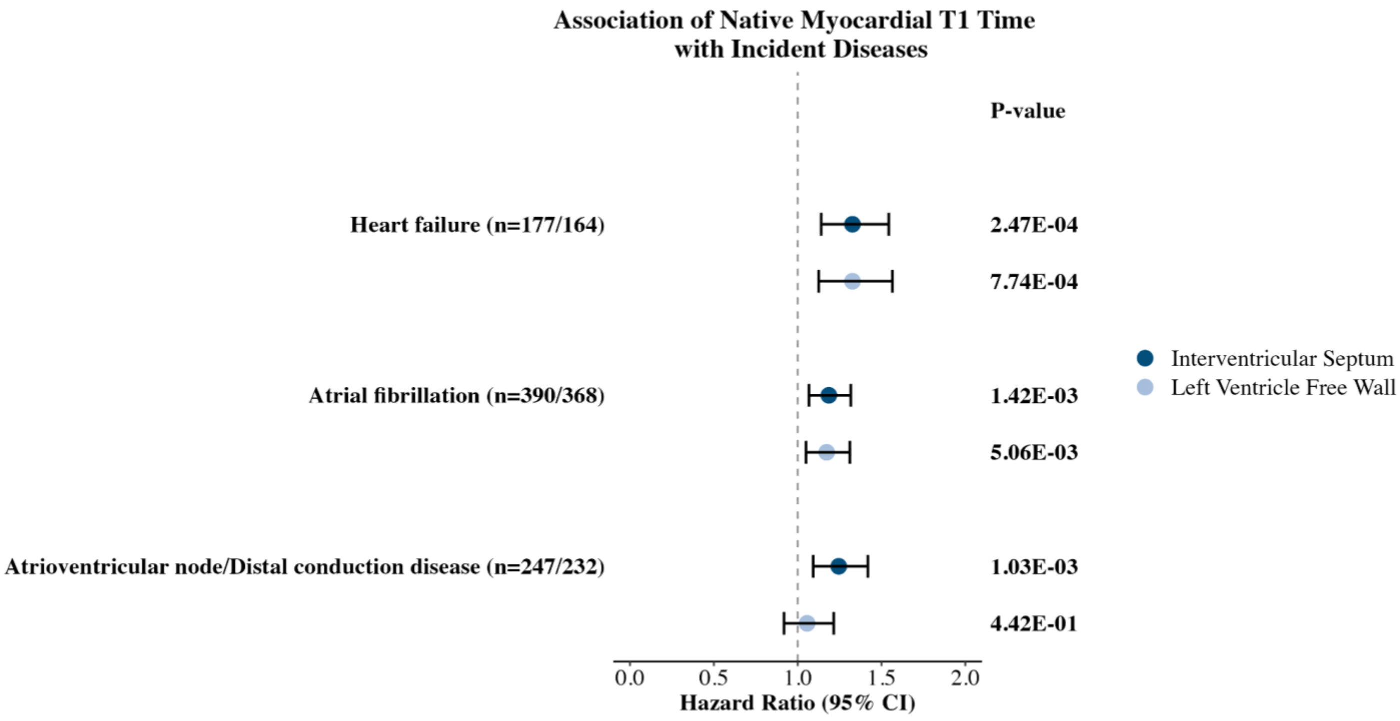

Supplemental Figure 10.

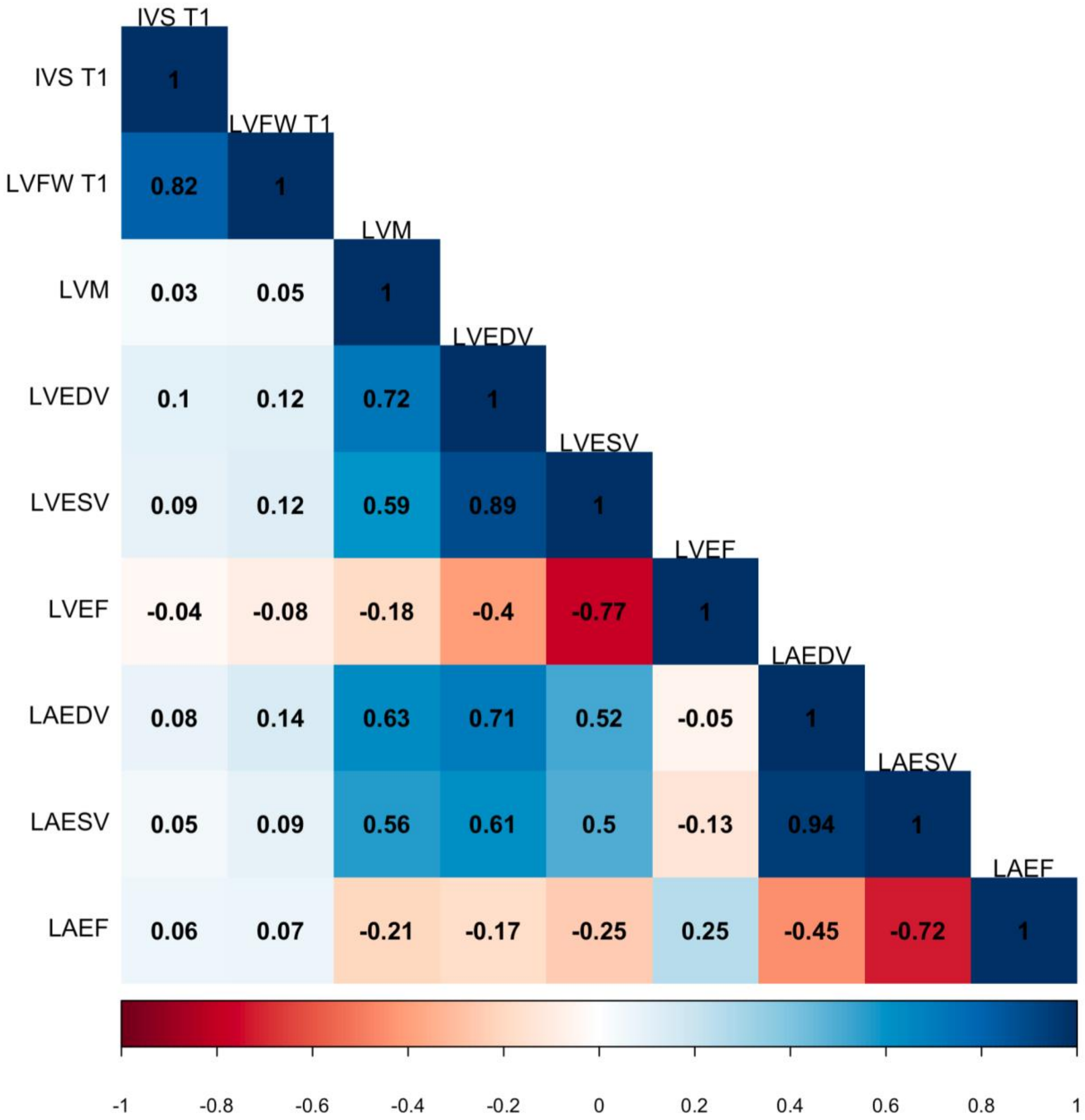

Supplemental Figure 11.

a

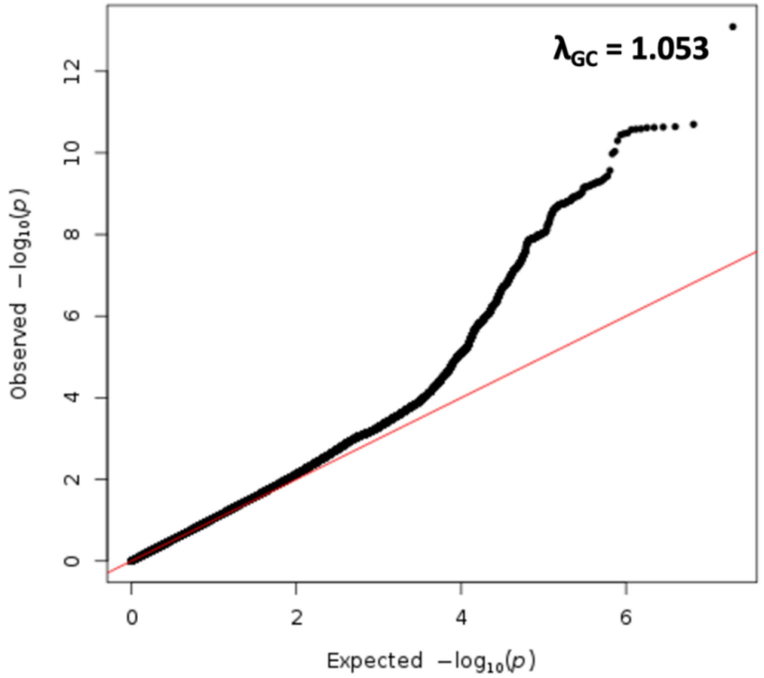

b

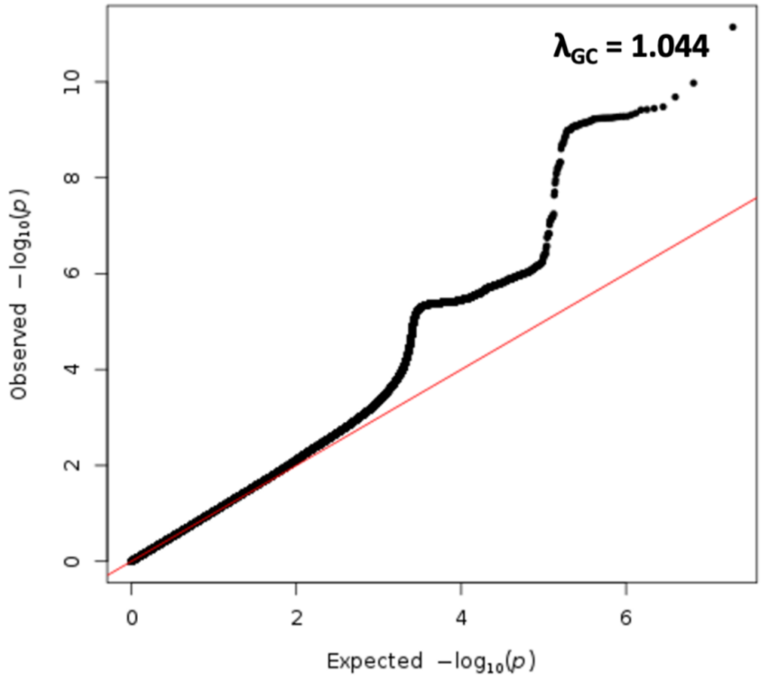

Supplemental Figure 12.

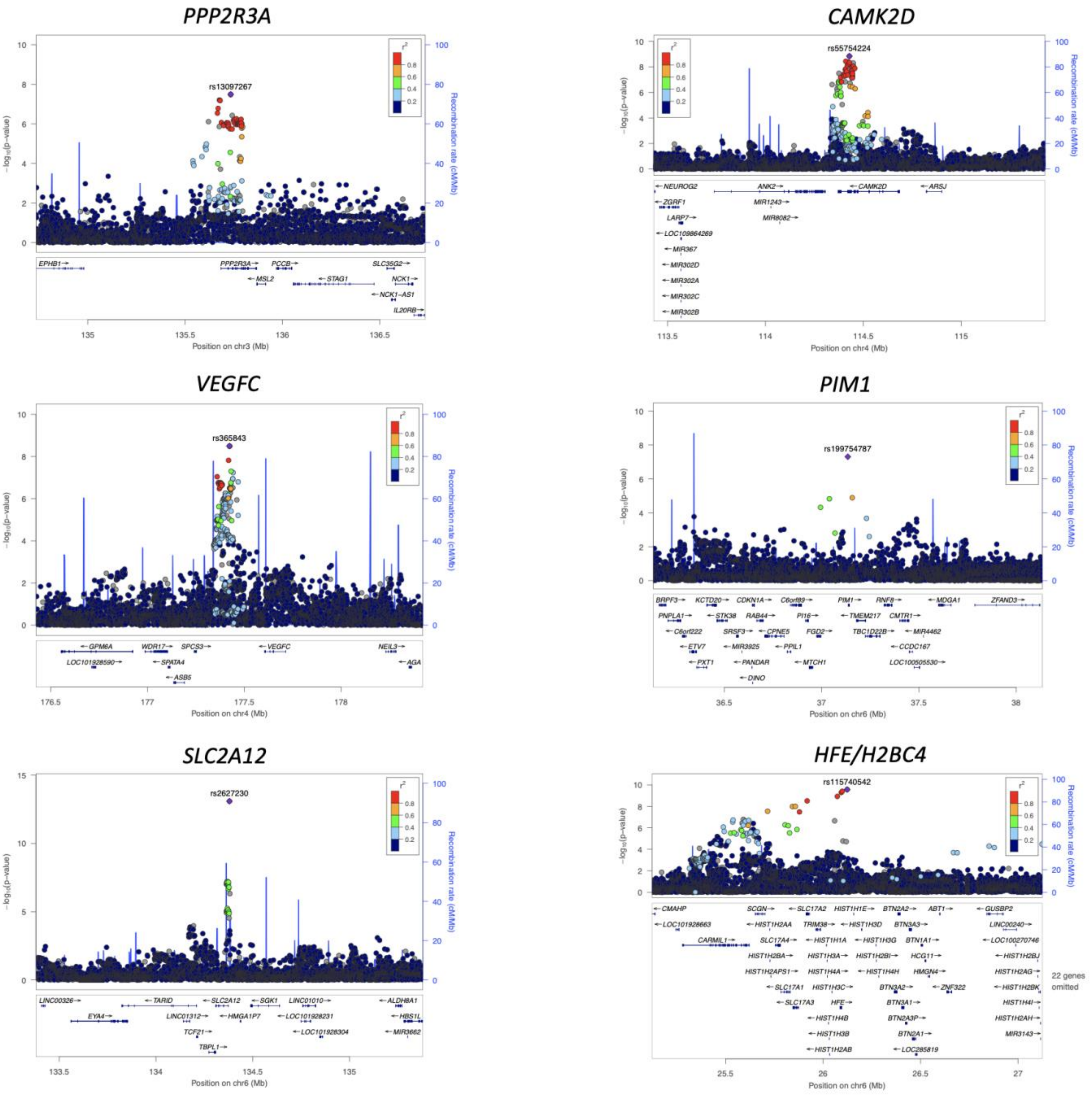

### SOD2

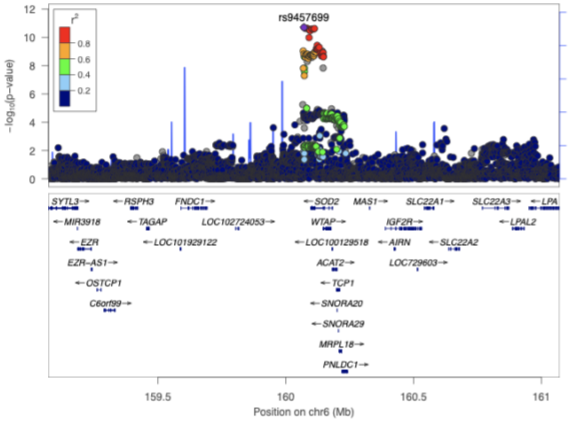

### KANK1

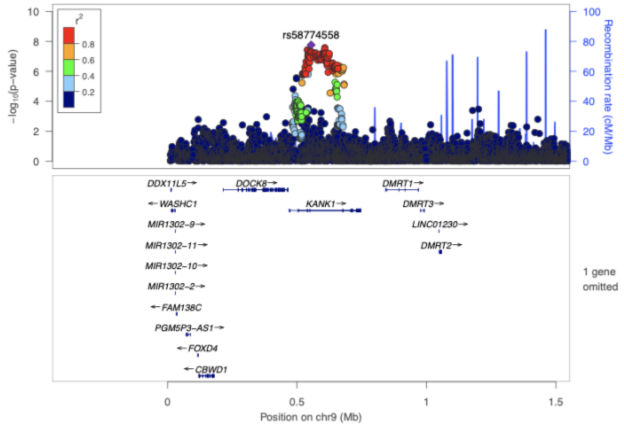

### ADAMTSL1

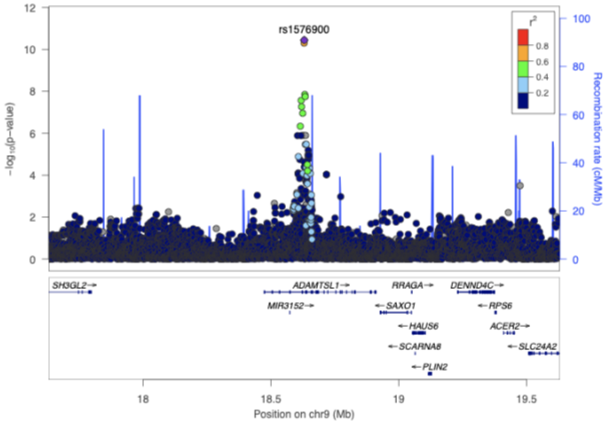

### MYH7B

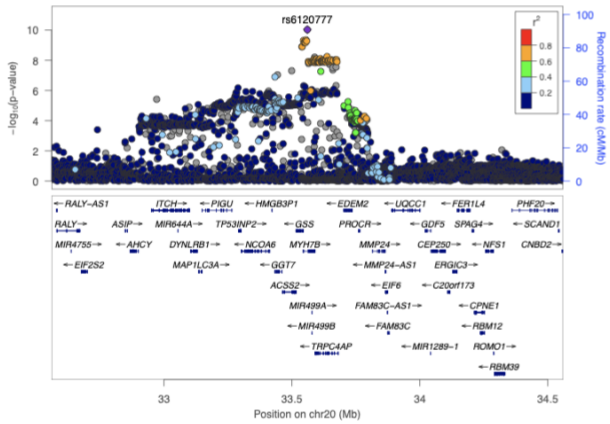

### TMPRSS6

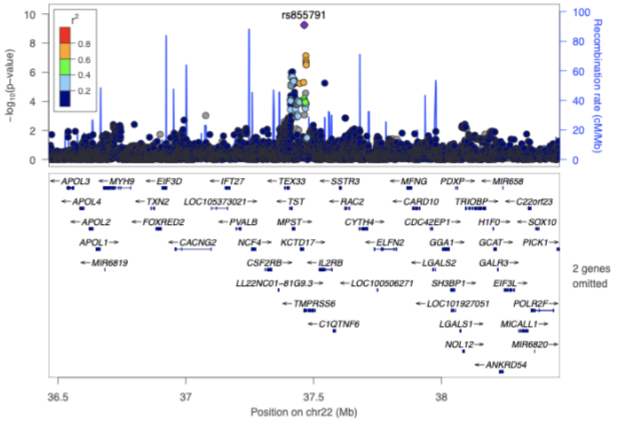

Supplemental Figure 13.

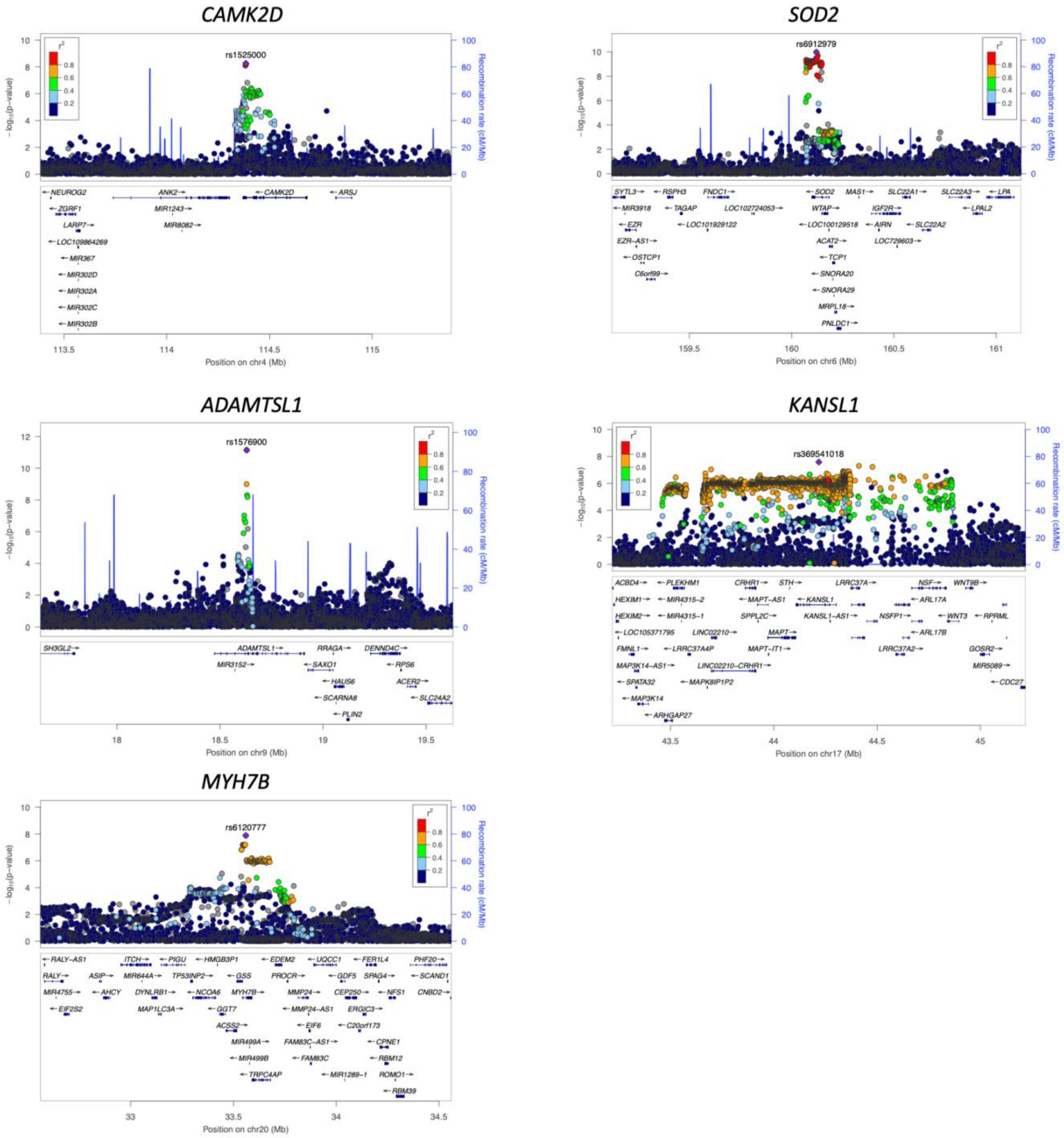

Supplemental Figure 14.

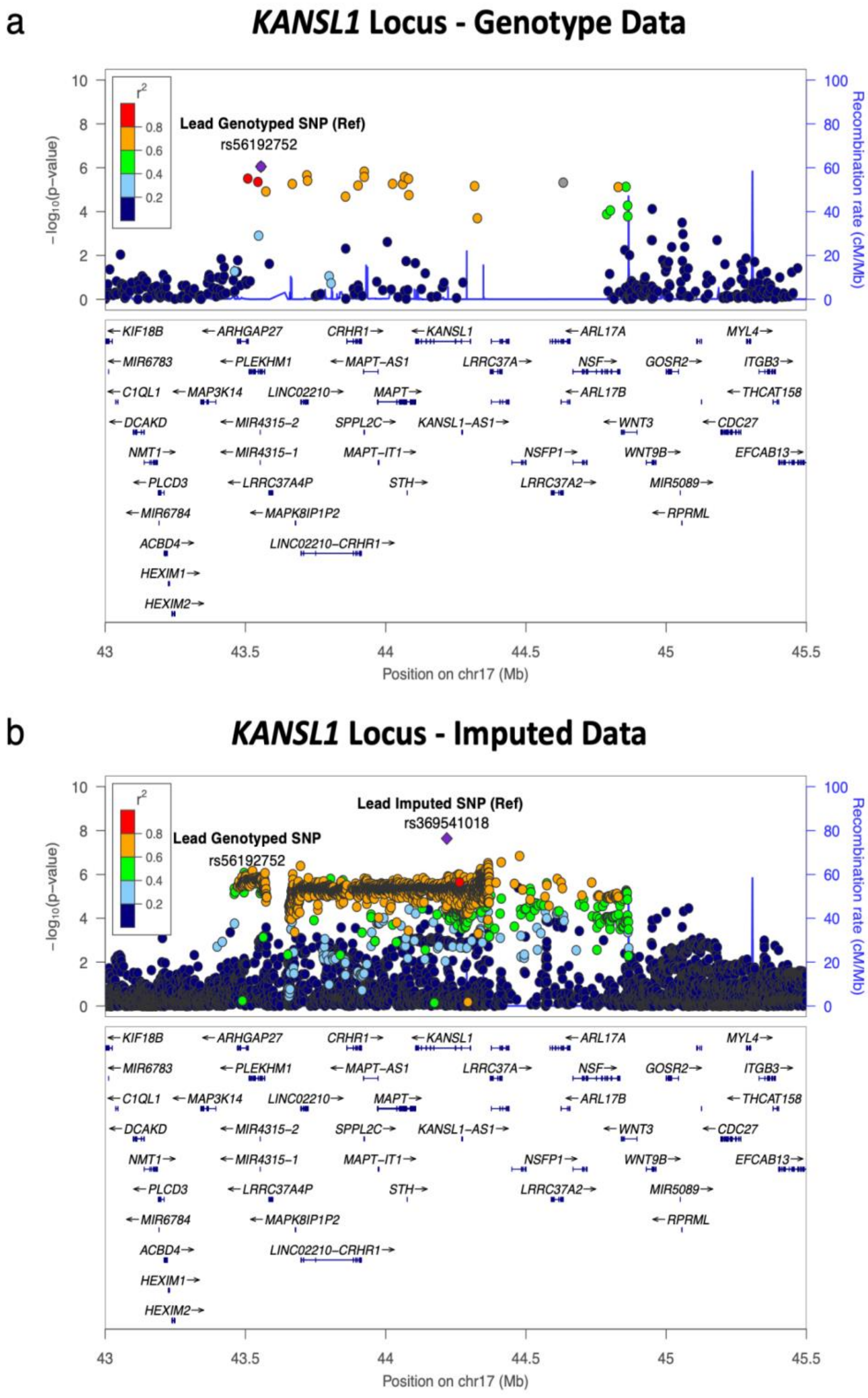

Supplemental Figure 15.

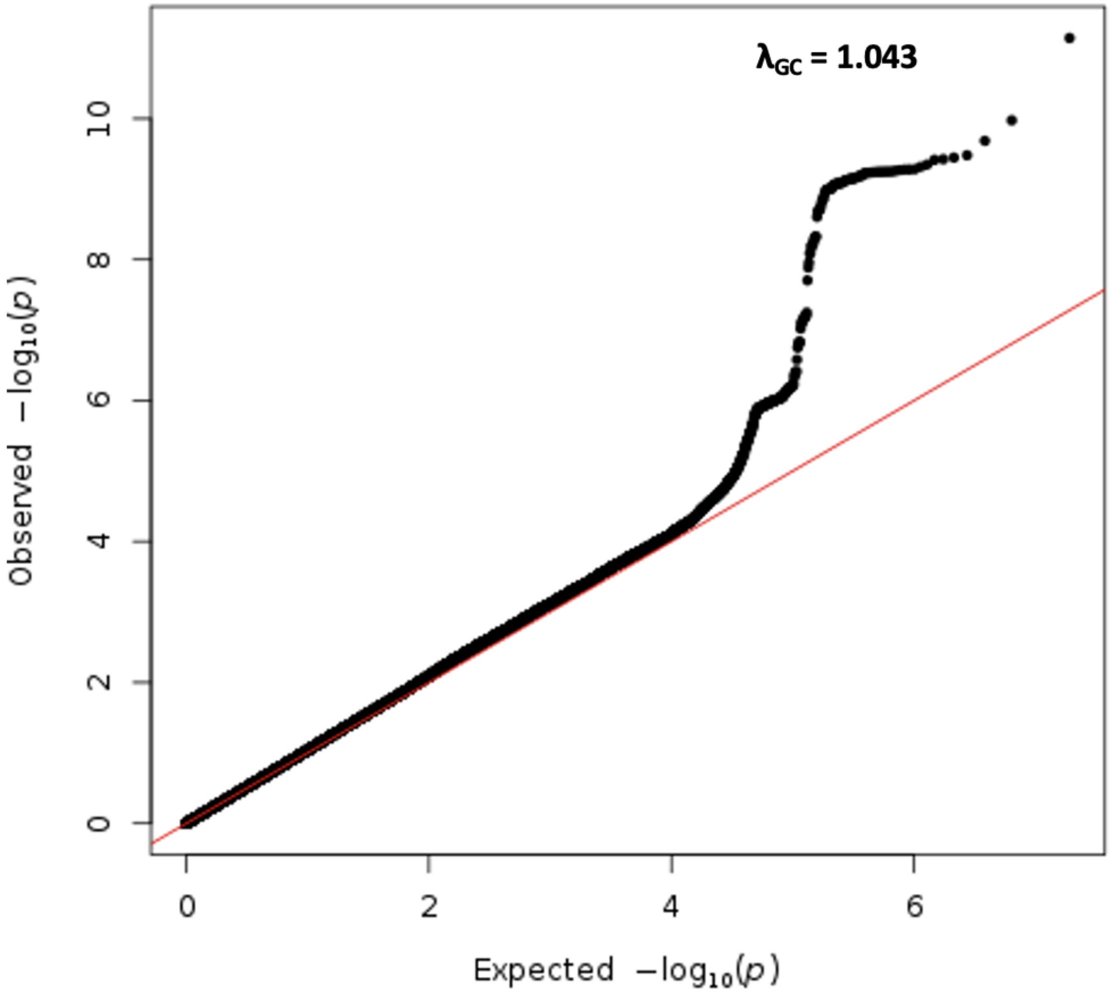

Supplemental Figure 16.

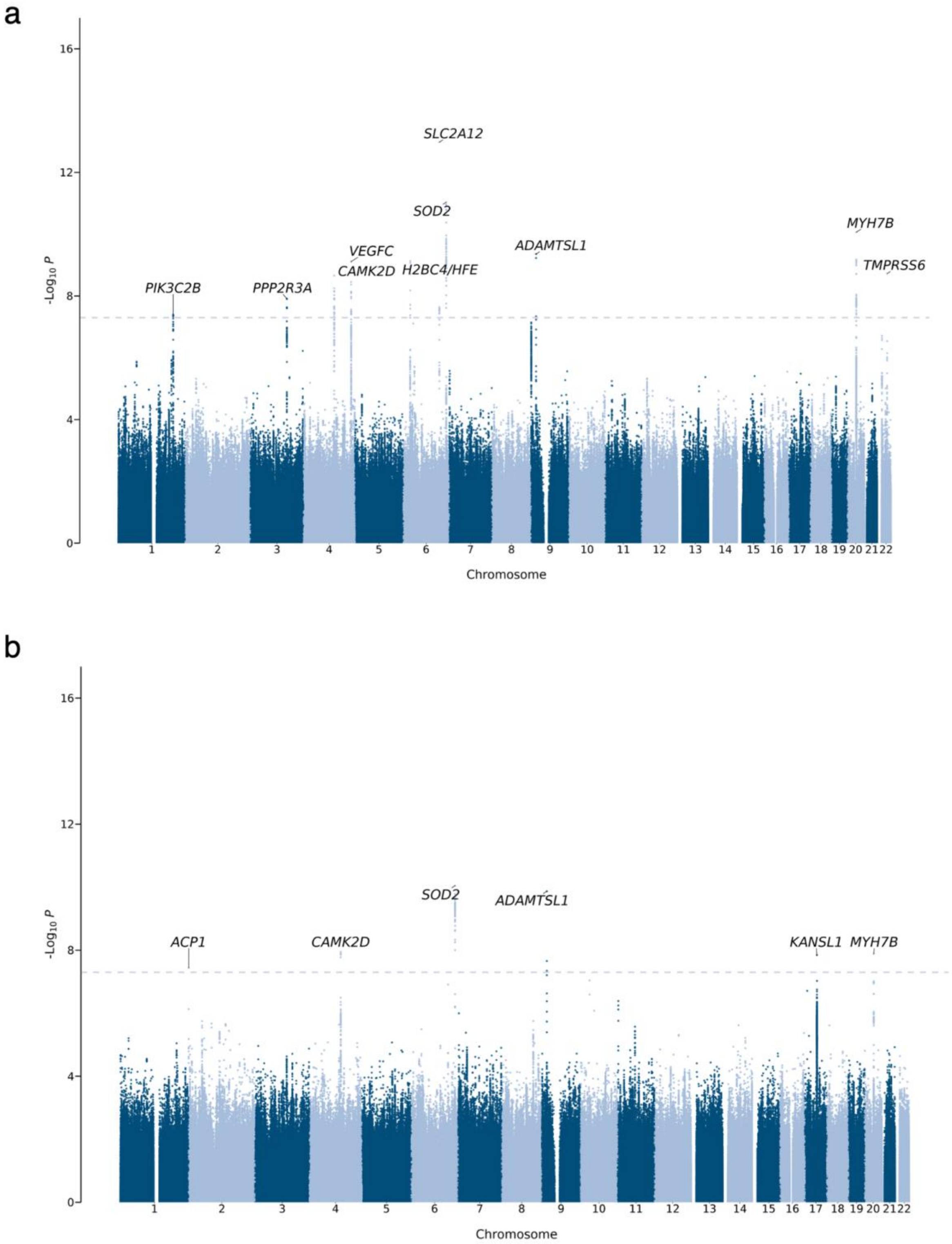

Supplemental Figure 17.

a

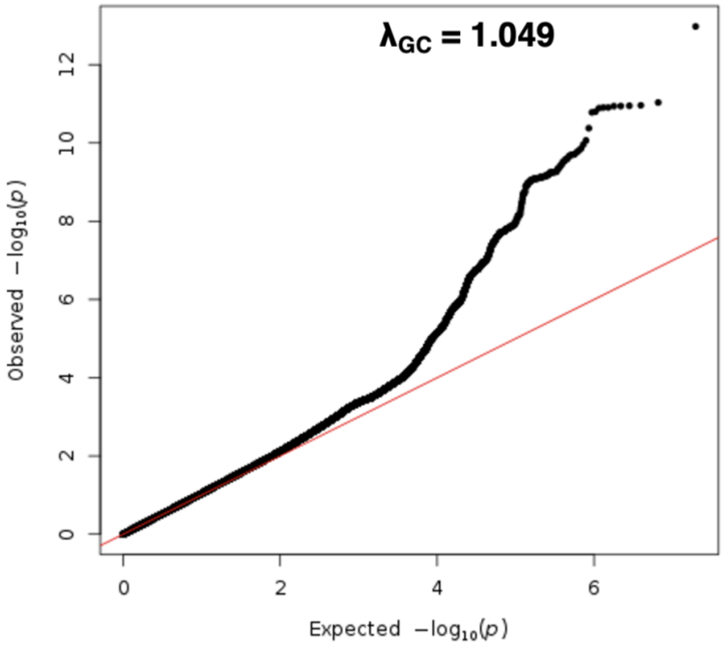

b

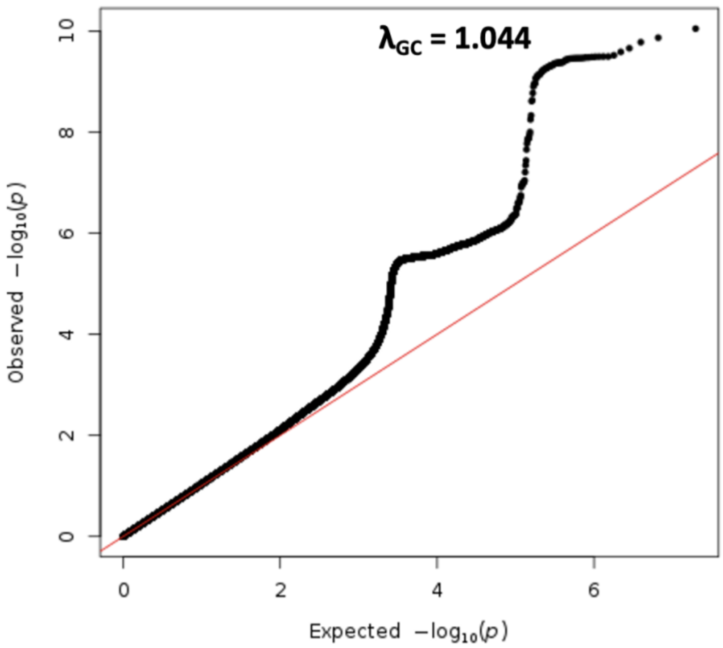

Supplemental Figure 18.

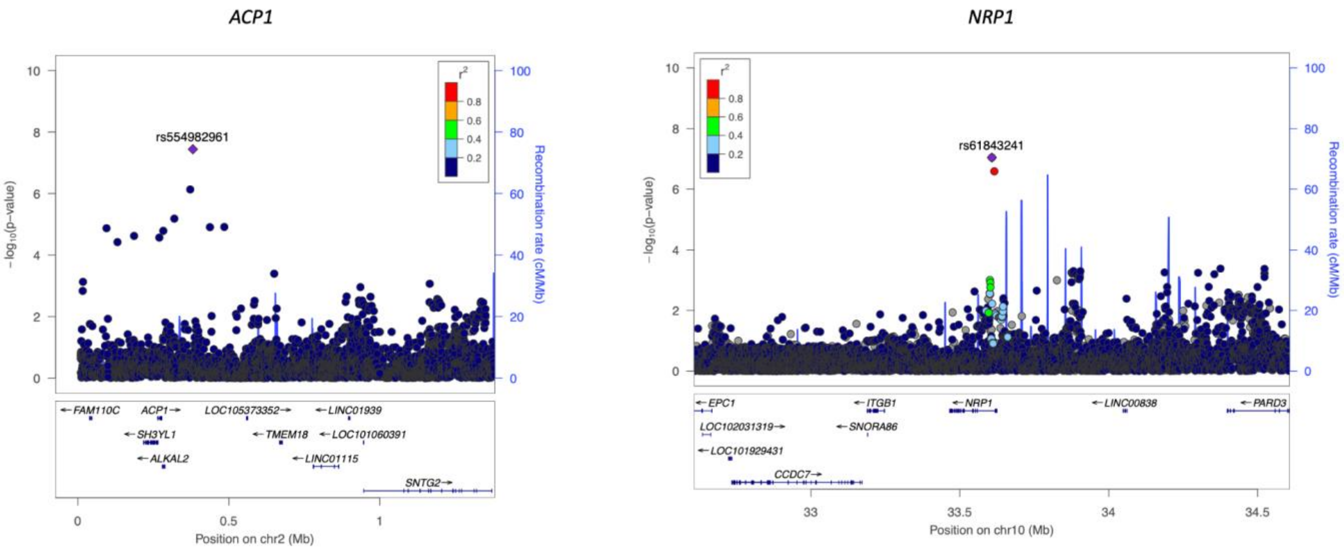

Supplemental Figure 19.

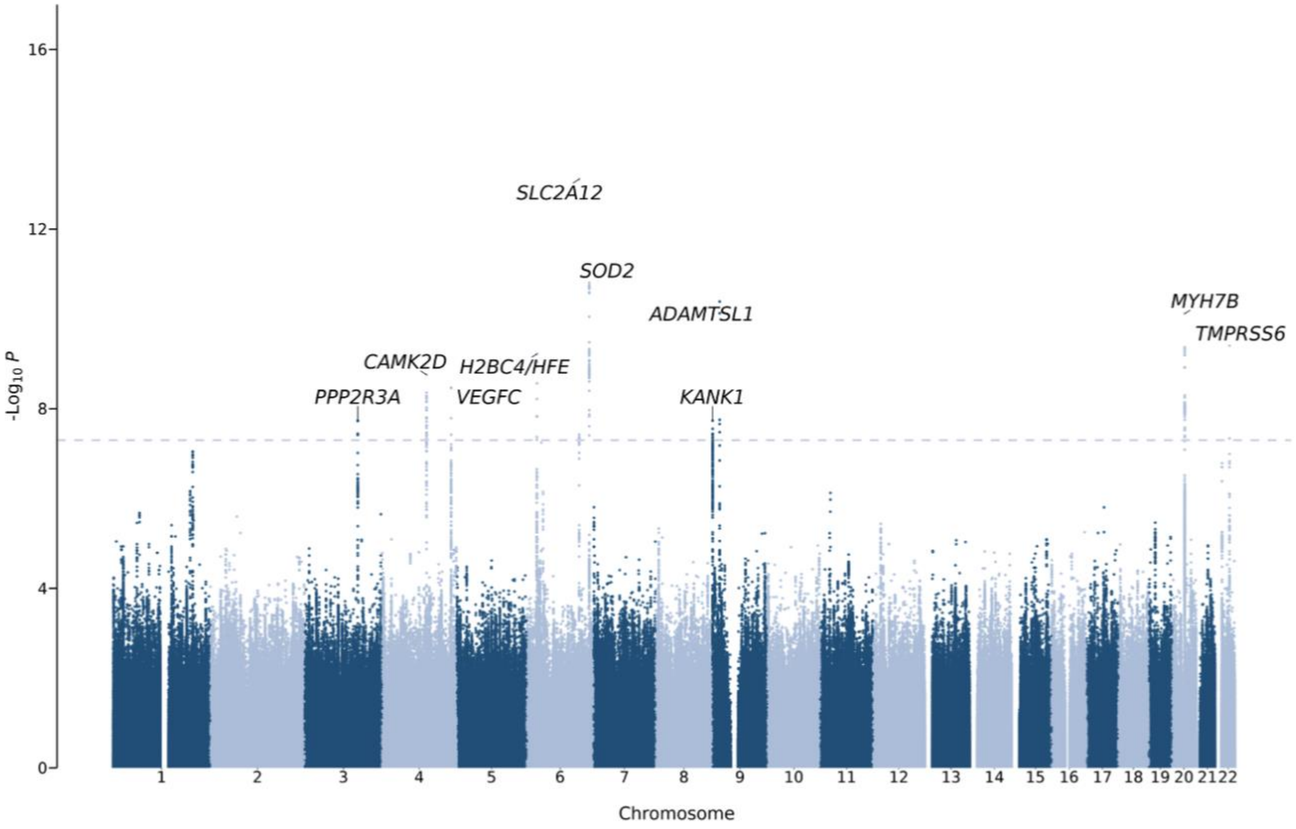

Supplemental Figure 20.

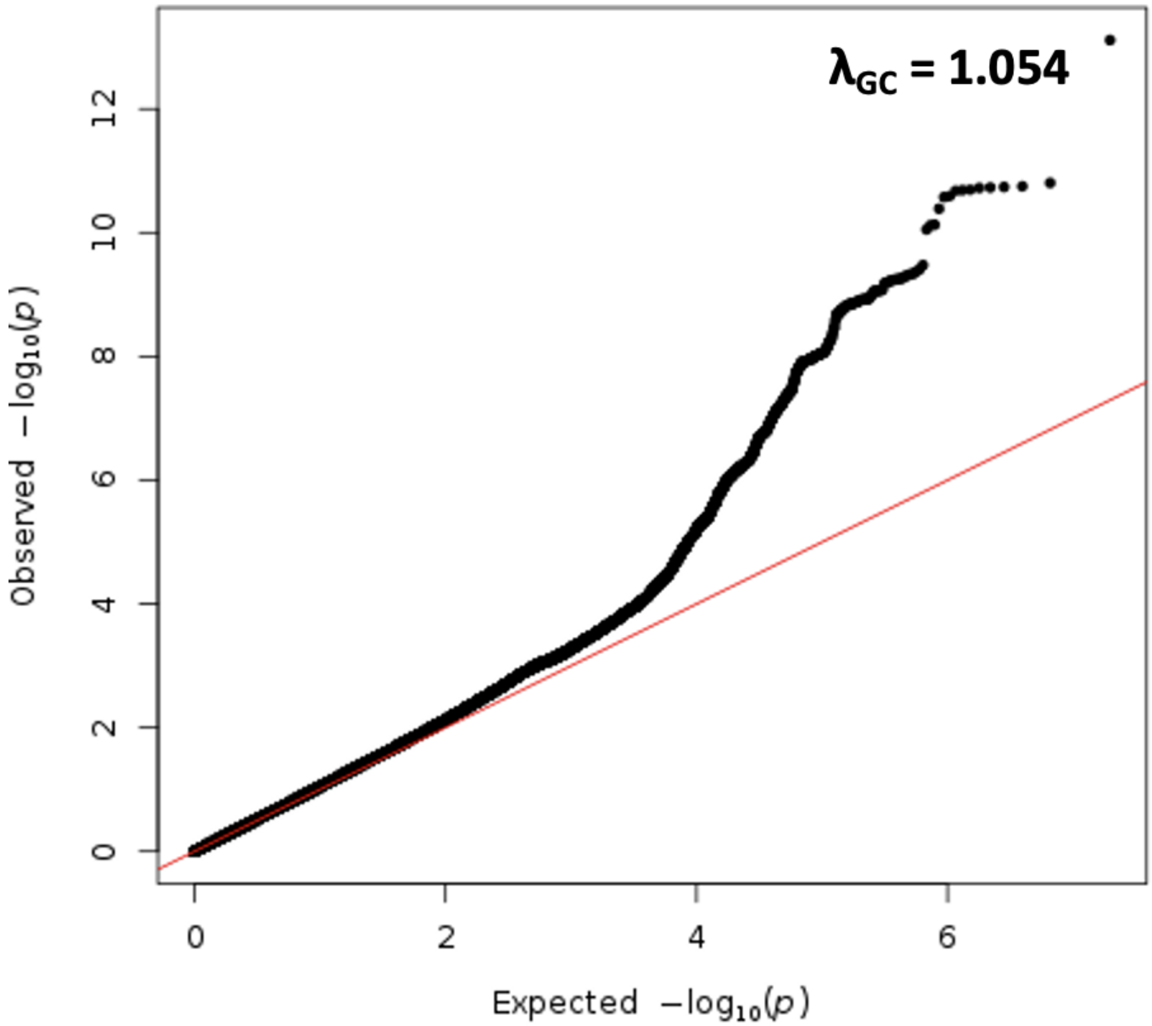

Supplemental Figure 21.

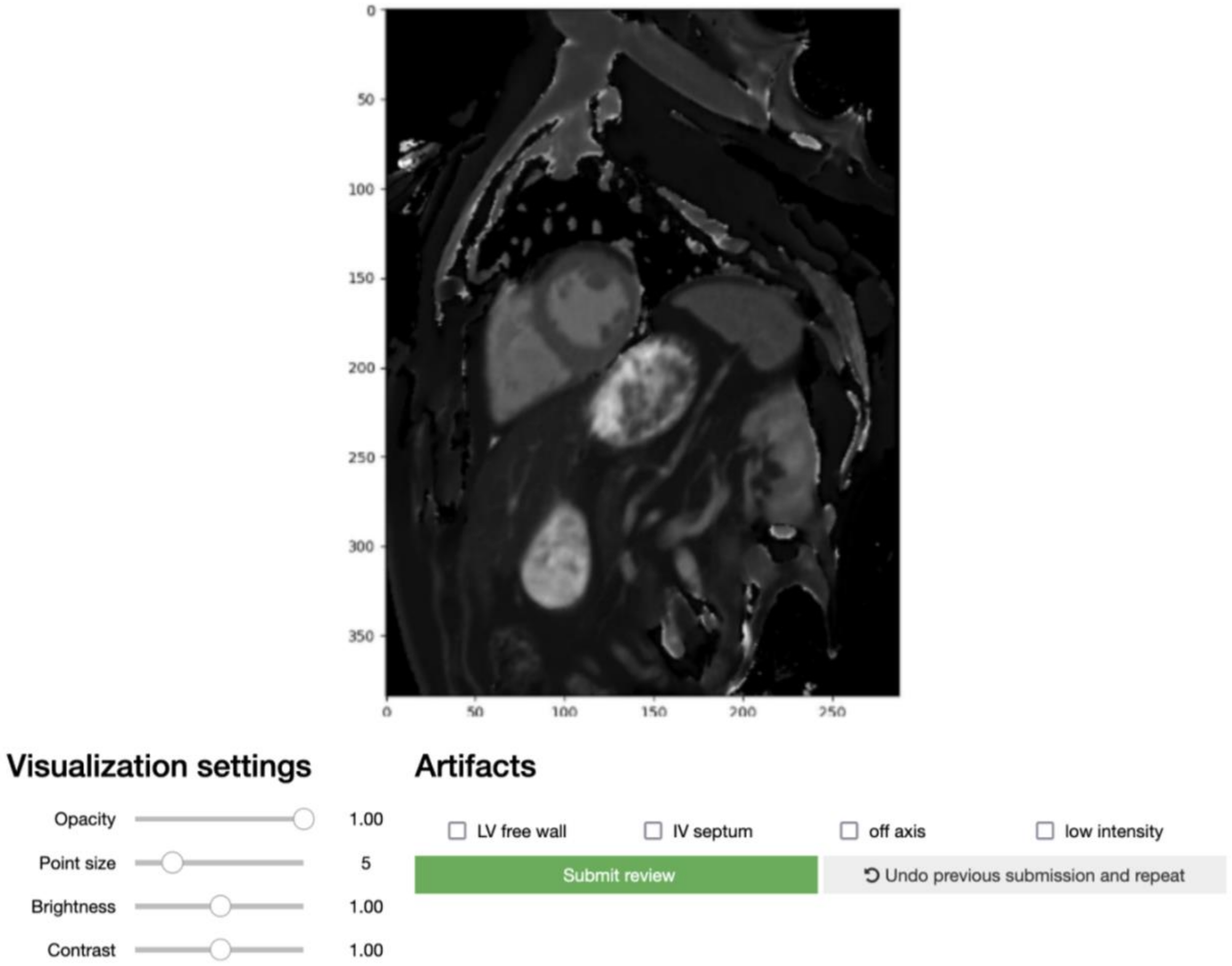

Supplemental Figure 22.

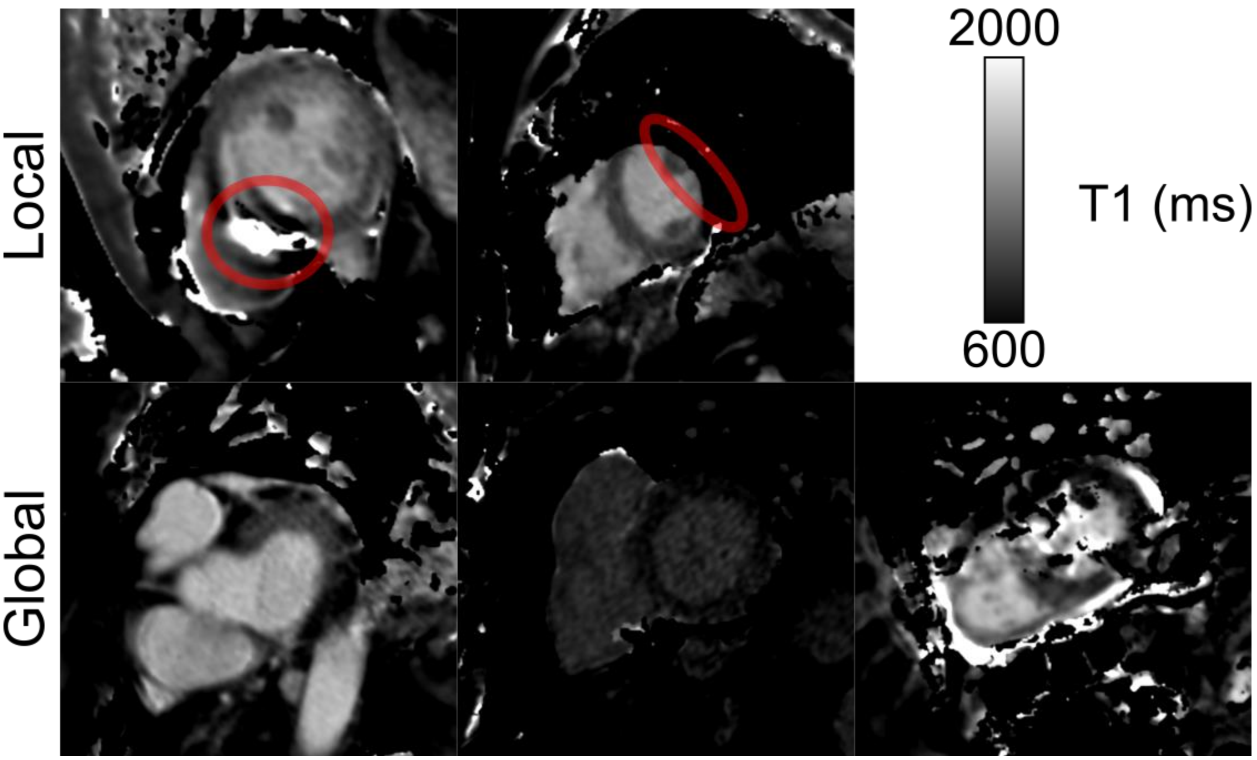
